# Prebiotic activity of lactulose optimizes gut metabolites and prevents systemic infection in liver disease patients

**DOI:** 10.1101/2023.02.14.23285927

**Authors:** Matthew A. Odenwald, Huaiying Lin, Christopher Lehmann, Nicholas P. Dylla, Ramanujam Ramanswamy, Angelica Moran, Alan L. Hutchison, Matthew R. Stutz, Mark Dela Cruz, Emerald Adler, Jaye Boissiere, Maryam Khalid, Jackelyn Cantoral, Fidel Haro, Rita A. Oliveira, Emily Waligurski, Thomas G. Cotter, Samuel H. Light, Kathleen G. Beavis, Anitha Sundararajan, Ashley M. Sidebottom, K. Gautham Reddy, Sonali Paul, Anjana Pilliai, Helen S. Te, Mary E. Rinella, Michael R. Charlton, Eric G. Pamer, Andrew I. Aronsohn

**Affiliations:** Department of Medicine, Section of Gastroenterology, Hepatology, and Nutrition, University of Chicago Medicine; Duchossois Family Institute, The University of Chicago; Department of Medicine, Section of Infectious Diseases, The University of Chicago; Department of Pathology, The University of Chicago; Department of Medicine, Division of Pulmonary and Critical Care Medicine, Cook County Health; Department of Medicine, Section of Cardiology, The University of Chicago; Division of Digestive and Liver Diseases, UT Southwestern Medical Center

## Abstract

Progression of chronic liver diseases is precipitated by hepatocyte loss, inflammation and fibrosis. This process results in the loss of critical hepatic functions, increasing morbidity and the risk of infection. Medical interventions that treat complications of hepatic failure, including antibiotic administration for systemic infections, impact gut microbiome composition and metabolite production. Using a multi-omics approach on 850 fecal samples from 263 patients with acute or chronic liver disease, we demonstrate that patients hospitalized for liver disease have reduced microbiome diversity and a paucity of bioactive metabolites. We find that patients treated with the orally administered but non-absorbable disaccharide lactulose have increased densities of intestinal *Bifidobacteria* and reduced incidence of systemic infections and mortality. *Bifidobacteria* metabolize lactulose, produce high concentrations of acetate and acidify the gut lumen, which, in combination, can reduce the growth of antibiotic-resistant pathobionts such as Vancomycin-resistant *Enterococcus faecium*. Our studies suggest that lactulose and *Bifidobacteria* serve as a synbiotic to reduce rates of infection in patients with severe liver disease.

## INTRODUCTION

The incidence of chronic liver disease (CLD) is rising, in part resulting from increased rates of metabolic syndrome and exposure to toxins, including alcohol^1–4^. Left untreated, all etiologies of CLD converge on the final common endpoint of cirrhosis with similar complications resulting from portal hypertension. CLD, in its early stages, can be clinically silent until hepatic decompensation, characterized by the development of ascites, variceal bleeding or hepatic encephalopathy (HE). Decompensating events often recur with increasing frequency, resulting in death or the need for liver transplant^5,6^. Treatment of advanced CLD is largely supportive and the ability to modify the clinical course of cirrhosis remains limited. Patients with decompensated cirrhosis frequently receive antibiotic treatment for HE, which is exacerbated by the activity of gut microbes and infection. The current first line treatment for HE is the non-absorbable disaccharide lactulose^7^. The mechanism through which lactulose reduces serum ammonia levels is incompletely known, and its role as a prebiotic that modifies the microbiome remains controversial ^8–12^. Moreover, there are varying reports illustrating the effects of lactulose on development of systemic infections and other complications of CLD, including death^13,14^. Together, these reports indicate that further evaluation is needed to fully understand the effects of lactulose on gut microbiome compositions and function as well as important clinical outcomes in patients with liver disease, including development of systemic infections.

The gut microbiome impacts human health and a wide range of diseases. Given the bidirectional anatomic communication between the liver and the gut via the portal vein and biliary tree, it is postulated that the microbiome plays a role in liver disease pathogenesis. Preclinical studies implicate the gut microbiome as a potential driver of non-alcoholic fatty liver disease (NAFLD) and alcoholic liver disease (ALD)^15–17^. While robust clinical evidence linking the microbiome to progression of liver disease is lacking, multiple observational studies have reported various gut microbiome “signatures” of advanced fibrosis and cirrhosis^9,18–21^.

Observational studies have also associated gut microbiome compositions with complications of end stage liver disease, including death^9,16,22,23^. Different commensal taxa are implicated in these studies, but in general, patients with higher burdens of potentially pathogenic taxa (e.g. *Proteobacteria* or *Enterococcus*) and lower prevalence of obligate anaerobic commensals (e.g. *Lachnospiraceae* and *Ruminococcus*) have a poor prognosis. This general principle is consistent with studies in non-hepatic diseases as well, including for patients undergoing bone marrow transplantation and patients with respiratory failure from COVID-19^24–26^.

While some studies have identified a potential difference in microbiome composition, the nature of these changes and functional consequences through microbial-derived metabolites (i.e. “the metabolome”) remain unaddressed. Microbe-derived metabolites contribute to intestinal epithelial cell differentiation and barrier formation^27–29^ and also regulate mucosal innate and adaptive immune defenses ^26,30,31^. In the case of liver disease, fecal bile acid (BA) profiles have been correlated with progression of NAFLD to non-alcoholic steatohepatitis (NASH) and subsequently advanced fibrosis^32–34^. Additionally, both total serum bile acids and specific immunomodulatory circulating BA have been implicated in the progression and prognosis of liver disease^35–37^. While recent studies have identified bacterial species that generate health promoting metabolites, whether these reduce or enhance progression of chronic liver disease remains largely unexplored territory.

Systemic infections precipitate hepatic decompensation and contribute to morbidity and mortality in liver disease patients^38,39^. The gut microbiome has been implicated in the development of systemic infections in many disease states^24,40^, including cirrhosis^41^. A recent prospective study used untargeted serum metabolites and fecal 16S rRNA microbiota data to predict nosocomial infections^42^; however, many of the serum metabolites of interest were not microbially derived and did not substantially improve the ability to predict infection development beyond standard clinical metrics (including known infection, model of end stage liver disease (MELD) score, and leukocyte count)^42^. Antibiotics, while highly effective for treatment and prophylaxis against common infections in liver disease^43,44^, are associated with increasing antibiotic resistance genes in the gut microbiomes and subsequent poor clinical outcomes^45–48^. Small studies of probiotics and fecal microbiota transfer (FMT) have been performed primarily as a means to control HE with overall positive clinical results including improved cognitive tests and fewer episodes of overt HE ^49–53^. However, while generally safe, there are reports of FMT-mediated transmission of toxin-producing and drug-resistant *E. coli* to patients, including one treated for HE^54,55^. Thus, FMT has not been adopted as a current standard of care^56^.

To better understand the role of the gut microbiome in the development of CLD complications, including development of systemic infections, we performed a single center observational cohort study of hospitalized patients with liver disease. Metagenomic and metabolomic profiles of fecal samples obtained from hospitalized patients were correlated with liver disease outcomes. We demonstrate that the commonly prescribed disaccharide lactulose preferentially expands *Bifidobacteria* and, in the absence of systemic antibiotic administration, results in a protective fecal metabolome. Functional *Bifidobacteria* expansion is associated with decreased abundance of antibiotic-resistant pathobionts and improved patient outcomes, including reduced incidence of systemic infections and prolonged survival in hospitalized patients. Our study provides novel insights into the mechanism by which lactulose impacts outcome of patients with CLD and provides a rationale for optimizing gut microbiome compositions and functions to minimize the complications of liver disease.

## METHODS

### Study Design

This was a prospective cohort study of consecutive hospitalized adult hepatology patients at a single institution from April 2021 to April 2022. Inclusion criteria were age ≥18 years, ability to provide informed consent (either themselves or by proxy if unable to provide consent), and being treated on the hepatology consult service. Subjects who were younger than 18 years, unable to provide consent, had prior solid organ transplant, or a prior colectomy were excluded. Patients were enrolled as soon as possible upon hospital admission, most within 48 hours. Samples were obtained under a protocol approved by the Institutional Review Board at the University of Chicago (IRB21-0327). Written informed consent was obtained from all participants or their surrogate decision makers.

### Specimen Collection and Analysis

Fecal samples were collected as soon as possible following study enrollment. If able to provide additional samples, fecal samples were collected approximately every 2 days during hospitalization. Samples were collected in a similar manner on re-hospitalization up until 1 year post-enrollment or until death or transplant. Demographic and clinical data were collected through review of the medical record and through the University of Chicago Center for Research Informatics (CRI). Clinical laboratory values, death and transplant information was obtained by chart review. Ascitic fluid infections without an evident intraabdominal source were all termed spontaneous bacterial peritonitis (SBP) and were diagnosed with the standard clinical definition of either polymorphonuclear cells (PMN) of 250cells/mm^3^ or greater or a positive ascites culture^43,44^. Cultures growing common contaminants (i.e. components of skin flora) were considered negative if PMN were < 250cells/mm^3^ and the clinical team did not treat for SBP. Samples were paired with the closest stool sample that was within 14 days prior to the ascitic sample or 3 days after the ascitic sample. If a sample was positive, all subsequent samples for the next 30 days were considered surveillance paracenteses and excluded from analysis. If a sample was negative, subsequent samples, which were typically done to for volume management, were also excluded. This approach mitigated observing the effects of targeted antibiotic use on the microbiome after diagnosis of SBP, over-representing a single patient for repeated negative paracenteses, and ensured that each analyzed stool sample was paired with only one ascites sample. Blood stream infection was defined as having a blood culture with bacterial or fungal growth. Skin contaminants were again excluded (i.e. considered “negative” cultures) if the clinical team did not treat for bacteremia. Blood cultures were paired to the closest stool sample within 30 days prior or 7 days after the blood culture. Blood cultures 30 days after a positive culture (i.e. “surveillance cultures”) were excluded from analysis, and blood cultures drawn within 30 days after a negative culture were also excluded as “duplicates.”

### Metagenomic Analyses

Fecal samples underwent shotgun DNA sequencing. After undergoing mechanical disruptions with a bead beater (BioSpec Product), samples were further purified with QIAamp mini spin columns (Qiagen). Purified DNA was quantified with a Qubit 2.0 fluorometer and sequenced on the Illumina HiSeq platform, producing around 7 to 8 million PE reads per sample with read length of 150 bp. Adapters were trimmed off from the raw reads, and their quality were assessed and controlled using Trimmomatic (v.0.39)^57^, then human genome were removed by kneaddata (v0.7.10, https://github.com/biobakery/kneaddata). Taxonomy was profiled using metaphlan4 using the resultant high-quality reads^58^. Microbial reads then were assembled using MEGAHIT (v1.2.9)^59^, genes are called by prodigal (https://github.com/hyattpd/Prodigal). In addition, high-quality reads are queried against genes of interest, such as virulence factors, cazymes, and antibiotic resistance genes, using DIAMOND (v2.0.4)^60^, and hits are filtered with threshold > 80% identity, > 80% protein coverage, then abundance is tabulated into counts per million or reads per million mapped reads (RPKM).

Alpha-diversity of fecal samples was estimated using the Inverse Simpson Index, while beta diversity was assessed by using taxumap (https://github.com/jsevo/taxumap). Metagenomic information is publicly available on NCBI under BioProject ID PRJNA912122.

### Mouse fecal DNA isolation

DNA was extracted using the QIAamp PowerFecal Pro DNA kit (Qiagen). Prior to extraction, samples were subjected to mechanical disruption using a bead beating method. Briefly, samples were suspended in a bead tube (Qiagen) along with lysis buffer and loaded on a bead mill homogenizer (Fisherbrand). Samples were then centrifuged, and supernatant was resuspended in a reagent that effectively removed inhibitors. DNA was then purified routinely using a spin column filter membrane and quantified using Qubit.

### 16S sequencing

V4-V5 region within 16S rRNA gene was amplified using universal bacterial primers – 563F (5’-nnnnnnnn-NNNNNNNNNNNN-AYTGGGYDTAAA-GNG-3’) and 926R (5’-nnnnnnnn-NNNNNNNNNNNN-CCGTCAATTYHT-TTRAGT-3’), where ‘N’ represents the barcodes, ‘n’ are additional nucleotides added to offset primer sequencing. Approximately ∼412bp region amplicons were then purified using a spin column-based method (Minelute,Qiagen), quantified, and pooled at equimolar concentrations. Illumina sequencing-compatible Unique Dual Index (UDI) adapters were ligated onto the pools using the QIAseq 1-step amplicon library kit (Qiagen). Library QC was performed using Qubit and Tapestation and sequenced on Illumina MiSeq platform to generate 2×250bp reads.

### 16S qPCR

Extracted DNA was diluted to 20ng/ul to ensure concentrations fell within measurable range. Degenerate primers were diluted to 5.5mM concentration. Primer sequences are as follows: 563F (5’-AYTGGGYDTAAAGNG-3’) and 926Rb (5’-CCGTCAATTYHTTTRAGT-3’). Standard curves were generates using linearized TOPO pcr2.1TA vector (containing V4-V5 region of the 16S rRNA gene) transformed into DH5α competent bacterial cells. Five-fold serial dilution was performed on the purified plasmid from 10^8^ to 10^3^ copies/□l per tube. PCR products were detected using PowerTrack SYBR Green Master Mix (A46109). qPCR was run on QuantStudio 6 Pro (Applied Biosystems) with the following cycling conditions: 95°C for 10 min, followed by 40 cycles of 95°C for 30 s, 52°C for 30s, and 72°C for 1min. Copy numbers for samples were calculated using the Design and Analysis v2 software

### Metabolomic Analyses

Short chain fatty acids (SCFA, i.e. butyrate, acetate, propionate, and succinate) were derivatized with pentafluorobenzyl bromide (PFBBr) and analyzed via negative ion collision induced-gas chromatography-mass spectrometry ([–]CI-GC-MS, Agilent 8890)^61^. Eight bile acids (BA, i.e. primary: cholic acid; conjugated primary: glycocholic acid, taurocholic acid; secondary: deoxycholic acid, lithocholic acid [LCA], isodeoxycholic acid; modified secondary: alloisolithocholic acid [alloisoLCA] and 3-oxolithocholic acid [3-oxoLCA]) were quantified (µg/mL) by negative mode liquid chromatography-electrospray ionization-quadrupole time-of-flight-MS ([–]LC-ESI-QTOF-MS, Agilent 6546). Eleven indole metabolites were quantified by UPLC-QqQ LC-MS. Eighty-five additional compounds were relatively quantified using normalized peak areas relative to internal standards. Data analysis was performed using MassHunter Quantitative Analysis software (version B.10, Agilent Technologies) and confirmed by comparison to authentic standards. Quantitative fecal metabolomic information paired to fecal metagenomic information is publicly available on NCBI under BioProject ID PRJNA912122. Raw data files are publicly available on MetaboLights project ID MTBLS7046.

### Bacterial culture

The *Bifidobacteria longum* strains MSK.11.12 and DFI.2.45 were previously derived from two distinct healthy donor stool samples and whole genome sequenced (BioSample ID: SAMN19731851 and SAMN22167409). The vancomycin resistant *Enterococcus faecium* (VRE) strain used in this study was obtained from ATCC (strain 700221). Both bacterial strains were grown in anaerobic conditions in Brain-heart infusion broth (BHI broth, BD 237500). The pH was adjusted to 7.0 with NaOH. Media was supplemented with lactulose (Thermo Scientific, J60160-22) or sucrose (Fisher BP220-1) where indicated. For growth in media supplemented with short chain fatty acids, media was supplemented with either sodium butyrate (Sigma 303410), sodium succinate (Sigma S2378) or sodium acetate (Sigma S5636). For studies of bile acid metabolism, 10μg/mL of conjugated primary bile acid, either glycocholic acid (Sigma T4009) or taurocholic acid (EMD Millipore, 360512), was added to the media. All growth curves were obtained in anaerobic culture conditions at 37°C on a BioTek EPOCH2 microplate reader with BioTek Gen 5 3.11 software. Growth curves were analyzed in GraphPad Prism Version 9.4.0. Lactulose concentrations in were measured using the EnzyChrom™ Lactulose Assay Kit (ELTL-100).

### Mouse studies

All mouse studies were approved by The University of Chicago Institutional Animal Care and Use Committee (IACUC, Protocol 72599). For germ-free studies, 8-12 week-old male C57BL/6 mice were used. Mice were bred and raised in a germ-free isolator, and after removal from the germ-free isolator, mice were handled in a sterile manner and individually housed in sealed negative pressure (BCU) isolators. Mice were treated with either regular sterile water or sterile water supplemented with filter sterilized lactulose at a final concentration of 20g lactulose/L of water. *B. longum* was grown to steady state in BHIS, pelleted, and resuspended in an equal volume of PBS, and previously germ-free mice were gavaged with 200μL of a freshly prepared suspension on 3 consecutive days. Fecal pellets were collected prior to and after *B. longum* inoculation at the indicated timepoints for 16S rRNA metagenomic analysis and targeted metabolomic analysis.

### Data sharing

In addition to the repositories specified above, all raw data included in the manuscript is publicly available at: https://github.com/yingeddi2008/DFILiverDiseaseMicrobiome.

## RESULTS

### Patient recruitment and disease characteristics

We enrolled 356 hospitalized patients with liver disease from April 2021 through April 2022. Of these 356 patients, 263 (73.8%) produced 850 stool samples that were analyzed by shotgun metagenomics and targeted metabolomics. The demographics and admission disease characteristics of patients who produced samples are shown in Table 1. Notably, alcohol use was the most common etiology of liver disease and accounted for 51.0% of patients. Patients had a median model of end stage liver disease MELD-sodium (MELD-Na) score of 18.69 (IQR 10.61 – 27.88). Of the 263 patients included in the analysis, 183 (69.6%) had known clinically significant portal hypertension upon hospitalization, which was defined either by hepatic venous pressure gradient (HVPG) ≥ 10mmHg, characteristic findings on imaging (enlarged portal vein, intraabdominal varices, splenomegaly, or ascites), or clinical features of ascites, varices, or hepatic encephalopathy. Over half (137, 52.1%) of the enrolled patients had some form of end organ dysfunction present on admission. End organ dysfunction included neurologic dysfunction (grade III or higher HE), circulatory dysfunction (mean arterial pressure < 60mmHg or requiring vasoactive agents), respiratory dysfunction (new or increasing oxygen requirement), or renal dysfunction (acute kidney injury or new hemodialysis needs). Taken together, these enrollment demographics and baseline disease characteristics indicate that, on average, this patient cohort has advanced liver disease with a poor prognosis.

### Fecal microbiome and metabolite profiles of liver disease patients are highly variable

To determine the range of fecal microbiome compositions in patients with liver disease, we performed shotgun sequencing on DNA extracted from 850 fecal samples collected from 263 liver disease patients. We used MetaPhlan4 to assign taxonomic compositions and Bray-Curtis distance to assess microbial diversity within the resulting dataset (Figure 1A). Immediately apparent is the low taxonomic alpha diversity in the majority of fecal samples (inverse Simpson range: 1.00 – 36.41, mean = 6.22, median = 3.86) and the expansion of bacterial taxa belonging to the *Enterococcus* genus (forest green) and *Enterobacteriaceae* family (red), which are common hallmarks of dysbiosis and are similar to microbiome domination detected in patients undergoing allogeneic hematopoietic cell transplantation^24,62^. Another striking observation is the high number of samples with marked expansion of the genus *Bifidobacteria* (shown in purple), which is commonly reported in breast fed infants but rarely seen in adults, suggesting that this may be a liver disease-specific finding.

**Figure 1:**
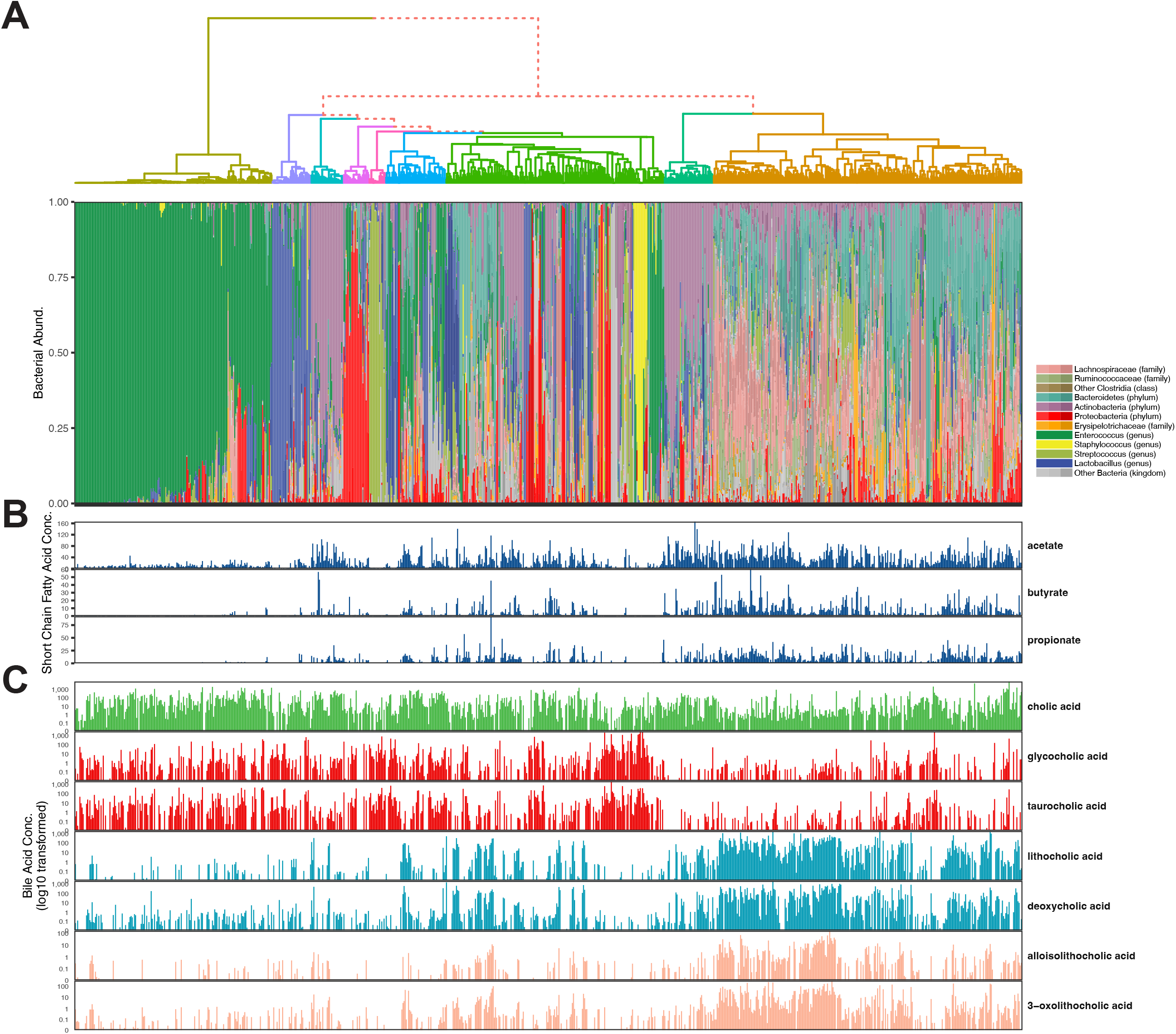
Fecal samples from hospitalized patients with liver disease display a wide range of microbiome compositions and metabolomic profiles. Hierarchical clustering of all 850 fecal samples is based on bray-curtis distance binned into nine clusters. (A) Relative taxa abundance by shotgun metagenomics is shown for each sample in the top panel, and (B and C) quantitative targeted metabolites for each corresponding sample are shown below. There is a wide variation in metagenomic compositions with some samples showing high biodiversity with anaerobic commensal bacteria compared to others that were dominated by a single taxon. (B) Short chain fatty acids and (C) both secondary and modified secondary bile acids are generally more abundant in the more biodiverse specimens. Units for SCFA are mM, and units for BA derivatives are in μg/mL.

While microbiome-derived or modified metabolites mediate many of the beneficial impacts on mucosal immune defenses and epithelial barrier functions, little is known about their production in liver disease patients harboring vastly different microbial populations. We used targeted GC- and LC-MS platforms to determine relative amounts 82 metabolites in all 850 fecal samples within our dataset, and the first sample from each patient is shown (Figure S1). Ranking of fecal samples from lowest to highest alpha diversity demonstrates the correlation between microbiome diversity and the relative amounts of SCFAs, branched chain FAs, aminated FAs, secondary bile acids, and various indole compounds and tryptophan metabolites. As expected, low diversity fecal samples have markedly reduced amounts of secondary bile acids but also reduced concentrations of SCFAs and a subset of indole compounds.

In order to obtain a more quantitative picture of fecal metabolite concentrations, we measured the concentration of SCFAs and primary and secondary bile acids in fecal samples from patients with liver disease. Despite some variation, higher proportions of commensal anaerobes and alpha-diversity generally coincided with greater levels of all measured short chain fatty acids (towards the right side of Figure 1B). Additionally, samples with more commensal anaerobes tended to have greater capacity to deconjugate primary bile acids and generate both secondary and modified secondary bile acids (Figure 1C). These findings demonstrate that the fecal microbiome and metabolome is highly variable in hospitalized patients with liver disease.

### Fecal metabolites correlate with expansion of distinct microbial taxa

To more precisely associate fecal metabolite concentrations with fecal microbiome compositions, fecal samples were assigned to 1 of 12 taxonomic groups on a taxonomic Uniform Manifold Approximation and Projection (taxUMAP) (Fig 2A). While all clusters displayed a wide range of short chain fatty acid production, our analysis presented in Figure 2 revealed that clusters that were characterized by samples with *Enterococcus* (forest green), *Proteobacteria* (red), or *Lactobacillus* (royal blue) as the most abundant taxa all had lower levels of short chain fatty acid production when compared to samples characterized by expansion of *Bifidobacteria* (purple), *Lachnospiraceae* (pink) or *Bacteroidetes* (aquamarine) (Fig 2A-D, S2A-C). On average, *Bifidobacteria*-expanded samples, which are prevalent in patients with liver disease, had higher acetate production (Fig 2B, S2A) but similar levels of butyrate (Fig 2C, S2B) and propionate (Fig 2D, S2C) compared to samples with high *Bacteroidetes* or *Lachnospiraceae* abundance. Consistent with a role for *Bifidobacteria* in acetate production, samples with greater than 10% *Bifidobacteria* abundance had a positive and statistically significant linear correlation with increasing acetate but not butyrate (Fig S2A-B, right-most panels). Propionate concentration negatively correlated with *Bifidobacteria* abundance (Fig S2C, right-most panel).

**Figure 2:**
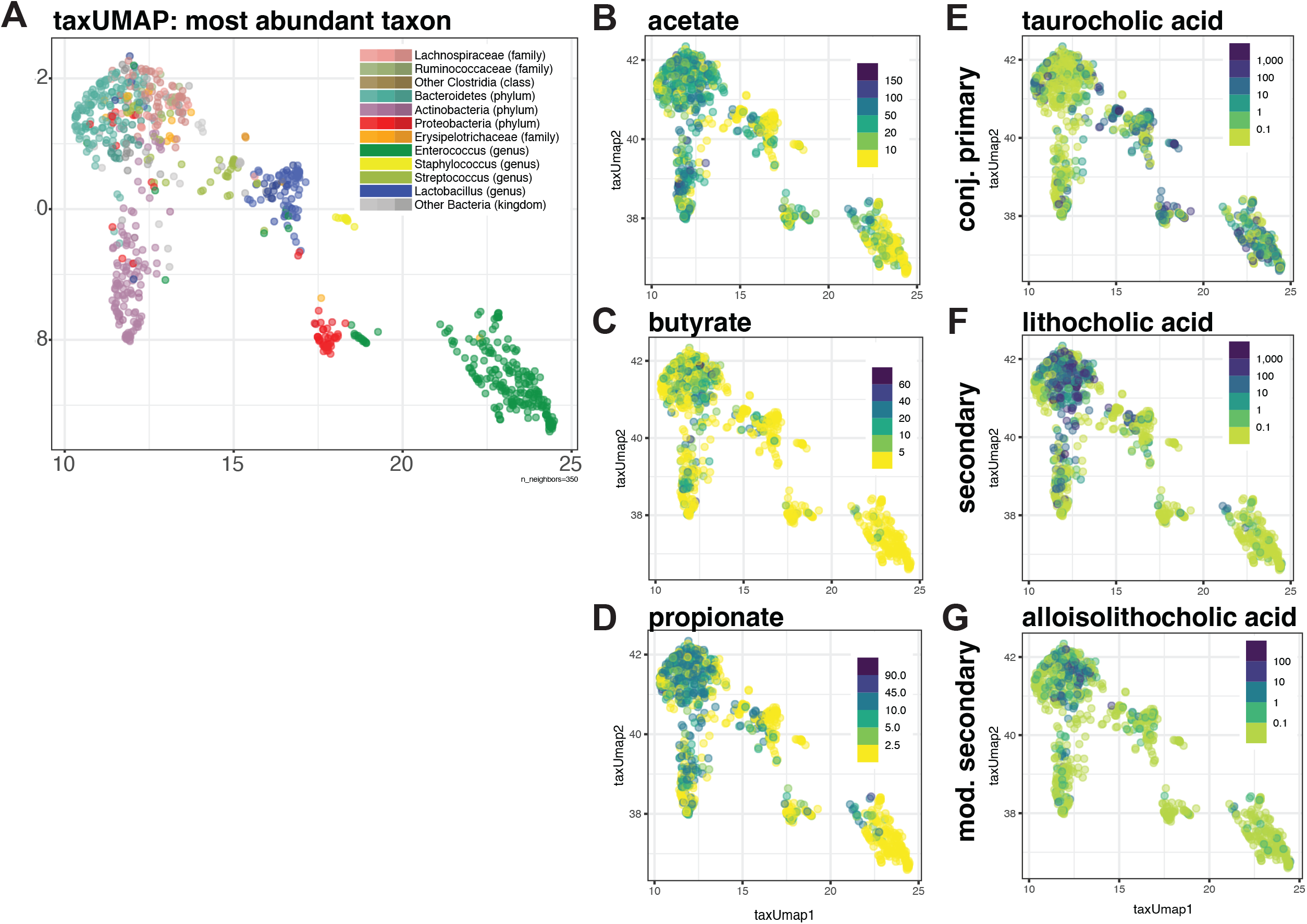
The unique *Bifidobacteria* expanded cluster has a distinct metabolite profile. (A) A taxUMAP was generated using 850 samples from 263 patients with liver disease. Each sample is represented by a single dot that is colored based on the most abundant taxon as indicated. Samples within the taxUMAP were pseudocolored based on the indicated (B-D) short chain fatty acid (SCFA) or (E-G) bile acid concentrations. Units for SCFA are mM, and units for BA derivatives are in μg/mL. Samples dominated by potentially pathogenic taxa (i.e. *Enterococcus* and *Proteobacteria*) were deficient in SCFA generation and generation of secondary bile acids compared to those dominated by anaerobic species. The unique *Bifidobacteria* cluster was particularly adept at generating acetate but produced secondary bile acids less efficiently than other anaerobe-dominated samples.

While all groups had detectable concentrations of primary bile acids, *Enterococcus* dominated samples had the highest (Fig S3). In contrast, conjugated primary bile acid levels were lower (Fig 2E, S3) and secondary (Fig 2F, S3) and modified secondary BA (Fig 2G, S3) levels were higher in samples with high levels of obligate anaerobes, including *Bifidobacteria, Bacteroidetes*, and *Lachnospiraceae*. Samples dominated by *Bifidobacteria* had higher levels of primary bile acids and lower levels of secondary bile acids compared to *Bacteroidetes*- and *Lachnospiraceae*-dominated samples, suggesting Bifidobacteria provide increased bile acid hydrolase activity and/or reduced efficiency of primary BA to secondary BA conversion. Taken together, our results indicate that a subset of patients with liver disease are predisposed to *Bifidobacteria* expansion with resultant acetate production and conjugated bile salt hydrolysis with reduced efficiency of secondary bile acid generation.

### Lactulose administration is associated with increased Bifidobacteria abundance and reduced antibiotic-resistant pathogen density

As lactulose is a common therapy administered to liver disease patients and has been reported to act as a prebiotic, we next sought to address how it might be impacting microbiome composition. Fecal samples obtained from liver disease patients after lactulose administration (lactulose given within 7 days prior to fecal sample) were taxonomically diverse, with many having marked *Bifidobacteria* expansion. In many samples, *Bifidobacteria* were the predominant genus (Fig 3A, B; range: 0.0 – 94.7%, mean: 10.9%, stdev: 10.9%). This contrasted with fecal samples obtained from patients who were not treated with lactulose (Fig 3A, B; range: 0.0 – 50.1%, mean: 3.57%, stdev: 3.57%, p < 0.001). High abundances of potentially pathogenic bacteria including *Enterobacteriaceae* and *Enterococcus* were detected in lactulose treated and untreated patients (Fig 3A). *Enterococcus* expansion was largely restricted to low-*Bifidobacteria* abundance samples and was associated with the presence of the vancomycin-resistance gene *vanA*, a result that was more striking in samples obtained in the presence of lactulose (Fig 3C). These results suggest that lactulose-mediated *Bifidobacteria* expansion may limit the expansion of Vancomycin-resistant *Enterococcus* (VRE) in the gut. Alternatively, treatment with broad-spectrum antibiotics may reduce the density of *Bifidobacterium* species while also eliminating other commensal species mediating resistance to VRE in lactulose-treated patients (Fig 3B) ^24,63^. Although the osmotic laxative effect of lactulose may affect the fecal metagenomic composition, we were unable to detect an association between stool consistency and *Bifidobacteria* abundance (Supplemental Data).

**Figure 3:**
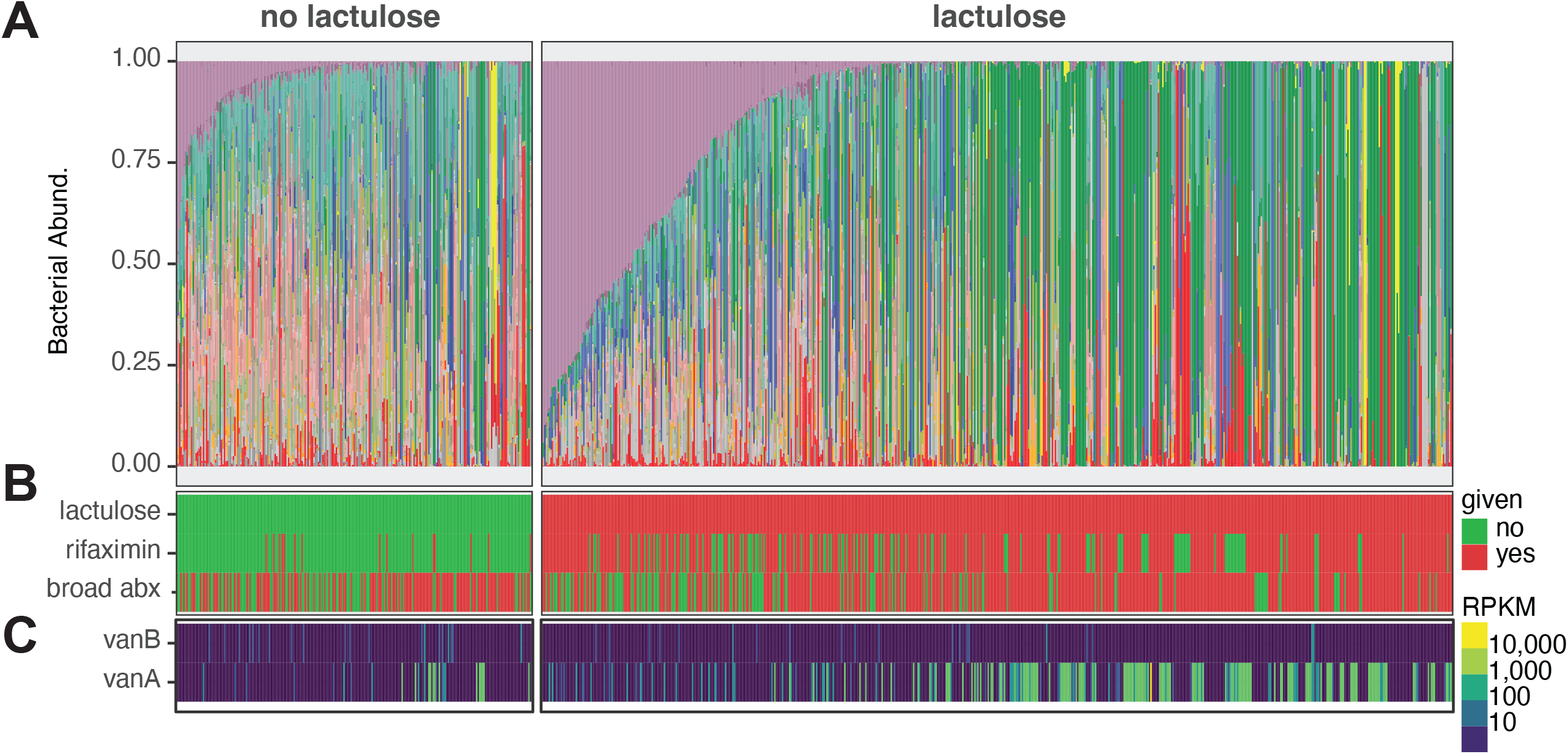
Lactulose use is associated with increased *Bifidobacteria* species abundance and reduced VRE abundance in the absence of systemic antibiotic use. (A) Relative abundance by shotgun taxonomy of all 850 samples is shown. Samples were arranged first by whether they were obtained without lactulose (left facet) or after lactulose administration (right facet) and then arranged by decreasing *Bifidobacterium* species (shown in purple) relative abundance from left to right. With decreasing *Bifidobacteria* abundance, there is higher abundance of *Enterococcus* (shown in forest green). (B) Medication information regarding lactulose, rifaximin, and broad-spectrum antibiotics is shown. Red indicates that a given treatment was administered prior to the sample being collected, and green indicates that a medication was not given prior to the sample being collected. (C) Relevant antibiotic resistance (*vanA* and *vanB*) genes were queried for each sample and are pseudocolored based on expression level normalized to gene length (RPKM).

### Lactulose-mediated Bifidobacteria expansion is associated with increased levels of multiple bioactive fecal metabolites

*Bifidobacteria* expansion, defined here as relative abundance ≥10%, was associated with marked changes in fecal metabolite profiles only in samples obtained in the presence of lactulose (Fig 4, S4). In lactulose-treated patients with *Bifidobacteria* expansion, the fecal metabolome was characterized by increased SCFA concentrations (including acetate, butyrate, and propionate), reduced conjugated primary bile acid concentrations (taurocholic, glycocholic, taurochenodeoxycholic, and glycochenodeoxycholic acids), increased secondary (lithocholic, isodeoxycholic acid) and modified secondary (alloisolithocholic, 3-oxo-lithocholic acid) bile acids, and indole metabolites (indole-3-lactic acid and indole-3-carboxaldehyde) (Fig 4A-C). Consistent with a known property of *Bifidobacteria* to hydrolyze conjugated primary bile acids, ratios of primary bile acids to conjugated primary bile acids were substantially higher in lactulose-treated samples with *Bifidobacteria* expansion (Fig 4D). While secondary bile acid levels were also higher in these samples, this was primarily due to an increased capacity to deconjugate primary bile acids rather than convert primary BA to secondary BA (Fig 4D, middle and right-most panels). In the absence of lactulose treatment, fecal samples with greater than 10% *Bifidobacteria* also had increased concentrations of SCFA (acetate and butyrate) and decreased conjugated primary bile acids (Fig S4). However, these changes were all modest compared to those from patients receiving lactulose, suggesting that lactulose impacts the metabolic activities of the gut microbiome in patients with liver disease (Fig S4).

**Figure 4:**
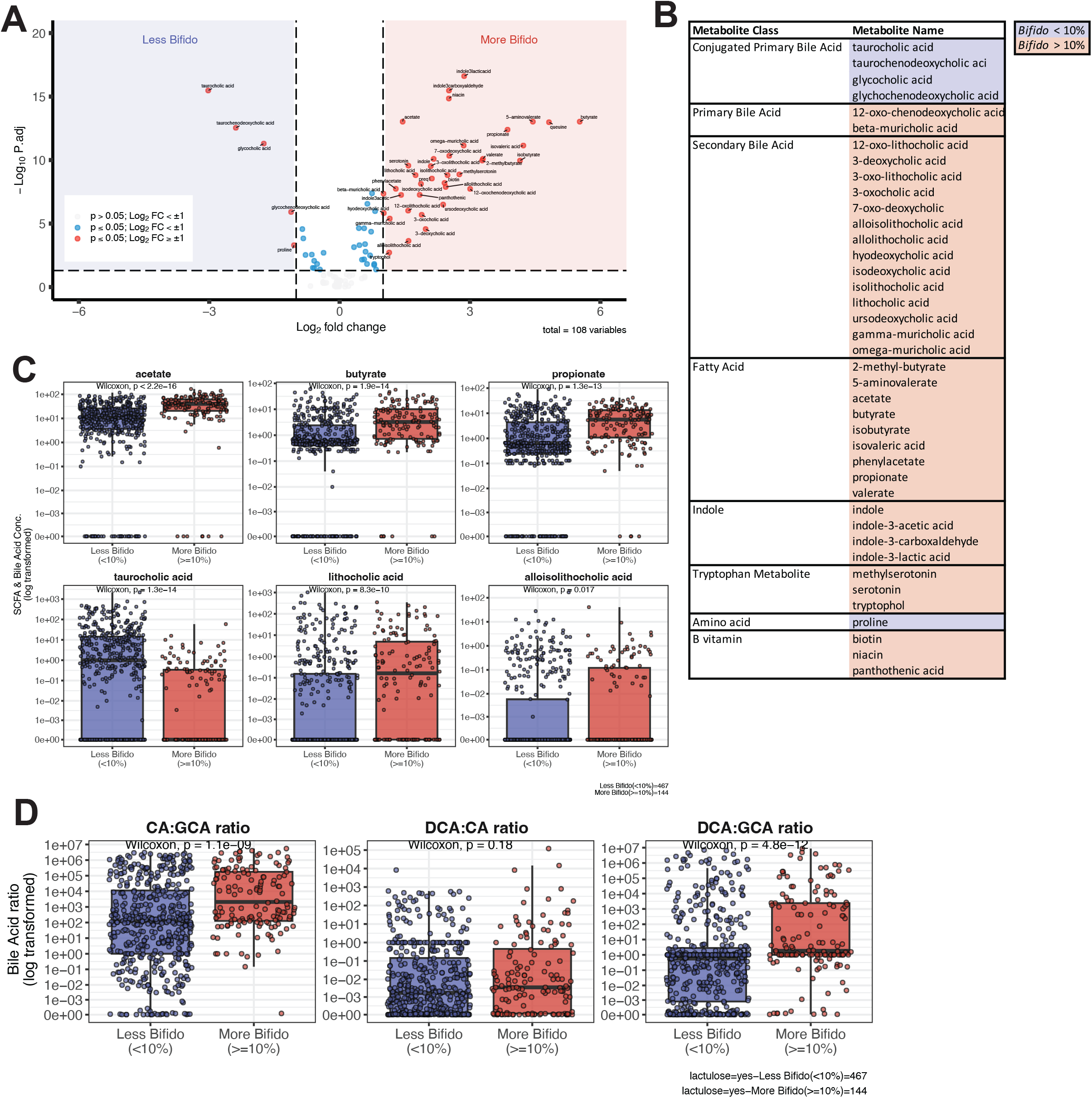
Lactulose-mediated *Bifidobacteria* expansion is associated with significant changes in bioactive fecal metabolites. (A) Volcano plot (log_2_fold change vs. log_10_p-value) of qualitative metabolites comparing samples with low (< 10%) vs. high (≥10%) *Bifidobacteria* abundance after lactulose exposure. P-values are corrected for multiple comparisons. Values with log2 fold-change > 1 (corresponding to a 2-fold change with a p-value < 0.05) were considered significant. (A and B) Notably, low *Bifidobacteria* abundance is associated with higher levels of unconjugated primary bile acids, whereas high *Bifidobacteria* abundance is associated with higher levels of beneficial bacterial metabolites, including short chain fatty acids, secondary and modified secondary bile acids, and immunomodulatory indole metabolites. (C) Bar graphs for select SCFA (acetate, butyrate, and propionate) and bile acids (taurocholic, lithocholic, and alloisolithocholic acid) where each point represents a single value. Median and interquartile range are indicated by the line and box, respectively. Statistical comparisons between groups were analyzed using the Wilcoxin rank sum. Units for SCFA are mM, and units for BA derivatives are in μg/mL. (D) Bile acid conversion from conjugated-primary BA to primary BA and then to secondary BAs was tested for each sample. Each point represents a ratio for an individual sample. Samples were grouped by whether they had expanded *Bifidobacteria* in the presence of lactulose. CA: cholic acid; GCA: glycocholic acid; DCA: deoxycholic acid.

### Lactulose enhances Bifidobacteria longum growth, acetate production and inhibition of VRE

Many fecal samples obtained from lactulose treated patients had high abundances of potentially pathogenic bacteria belonging to *Enterobacteriaceae* family (range: 0.0 – 99.2%, mean: 8.04%, median: 0.73%, stdev: 8.04%) and the *Enterococcus* genus (range: 0.0 – 100%, mean: 30.8%, median: 5.24%, stdev: 30.8%), with, on average, higher densities than those seen in fecal samples taken from patients not receiving lactulose (*Proteobacteria* mean: 6.38%, median: 1.25%; *Enterococcus* mean: 7.4%, median: 0.06%). While high abundances of *Proteobacteria* species were seen throughout a range of *Bifidobacteria* abundances, *Enterococcus* domination was seen predominantly in samples with reduced to absent *Bifidobacteria* abundance (Fig 3A, 5A).

**Figure 5:**
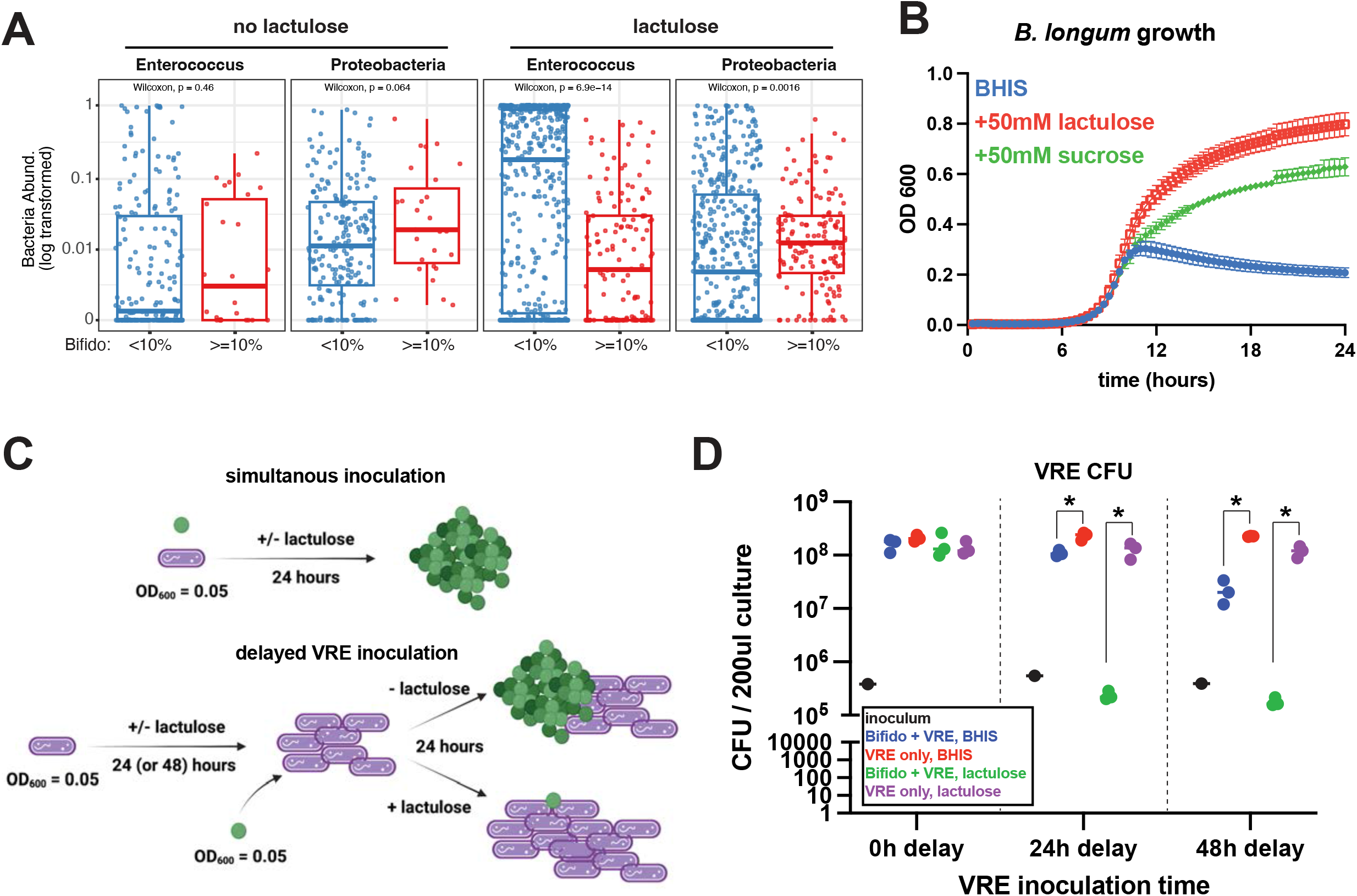
Lactulose-mediated *Bifidobacteria* expansion is associated with exclusion of antibiotic-resistant *Enterococcus* species. (A) Relative abundance of potentially pathogenic taxa *Enterococcus* and *Proteobacteria* were plotted based on lactulose exposure (no lactulose, left; lactulose, right) and relative abundance of *Bifidobacteria* (< 10%, blue; ≥10%, red). Neither taxa was changed at high or low *Bifidobacteria* abundance without lactulose, but with lactulose, *Enterococcus* abundance was significantly increased with low *Bifidobacteria* abundance in human fecal samples. *Proteobacteria* are modestly but statistically significantly increased in *Bifidobacteria* expanded samples from lactulose-treated patients. (B) A human-derived *B. longum* species was grown in BHIS media (blue) or BHIS supplemented with 50mM lactulose (red) or 50mM sucrose (green), and growth curves (OD_600_ over time) are shown. Growth curves are representative of three independent experiments done in at least triplicate. *B. longum* preferentially uses lactulose as an energy source. (C) Schematic of experimental design for *B. longum* and VRE co-culture. *B. longum* and VRE were grown to steady state and diluted to low density (OD_600_ = 0.05) prior to inoculation in either BHIS or BHIS supplemented with 50mM lactulose. Cultures were inoculated with both bacteria simultaneously (top) or *B. longum* was given either a 24-hour or 48-hour lead time prior to VRE inoculation (bottom). (D) After 24-hours of co-culture, serial dilutions were plated for VRE c.f.u. counts. Groups were compared using a two-tailed student’s t-test. The plot is from one experiment that is representative of three independent experiments done in at least triplicate.

To further investigate the inverse relationship between *Bifidobacteria* and VRE abundance, we isolated a *Bifidobacterium longum* strain from a healthy human donor and performed whole genome sequencing to demonstrate the presence of genes involved in lactulose metabolism and acetate production^64,65^. *B. longum* growth in culture was markedly augmented by addition of lactulose to media, and to a lesser extent by addition of sucrose (Fig 5B). Lactulose-induced growth augmentation was accompanied by increased media acidification and acetate, but not butyrate or propionate, production (Fig S5A, B). Similar to patient fecal samples dominated by *Bifidobacteria* and consistent with coding for bile salt hydrolase genes, monocultures of *B. longum* efficiently deconjugated conjugated primary bile acids but did not convert these to secondary bile acids (Fig S5C, D).

We next colonized germ-free (GF) mice with *B. longum* in the presence or absence of lactulose (Fig S6A). Lactulose was detectable in the stool of GF mice and led to increase stool water content (Fig S6B, C). *B. longum* efficiently colonized and persisted in the intestines of gnotobiotic mice independent of lactulose (Fig S6D). Consistent with *in vitro* functions, *B. longum*-colonized mice had measurable fecal acetate levels but did not generate butyrate or propionate (Fig S6E-G). Fecal acetate levels were similar between water and lactulose-treated mice at day 0, but on day 10 lactulose augmented acetate production (Fig S6E). On day 10 after *B. longum* colonization, fecal bile acid profiling demonstrated increased primary bile acids but undetectable secondary BA (Fig S6H-J).

To test whether *B. longum* inhibits VRE growth and to determine the impact of lactulose administration, we performed co-culture assays (Fig 5, C-D). While simultaneous inoculation of *B. longum* and VRE into media did not reduce VRE growth (Fig 5C-D), pre-culture of *B. longum* for 24h or 48h fully inhibited VRE growth in the presence of lactulose and only partially inhibited VRE growth in the absence of lactulose (Fig 5C-D). Serial dilutions of *B. longum* culture supernatant with fresh media sequentially reduced the inhibitory effect on VRE growth (Fig S7A, B), and alkalization of *B. longum* supernatant pH from 5.8 up to 7.2 restored VRE’s initial log phase growth (Fig S7C) but did not enable VRE to reach the maximal OD_600_ obtained in fresh, neutral pH media (Fig S7C). VRE growth is inhibited by addition of short chain fatty acids to the media, and inhibition is augmented at lower media pH (Fig S7D). Taken together, these data suggest that *B. longum* inhibits VRE growth through parallel mechanisms that include media acidification, SCFA production, and, to a lesser extent, nutrient deprivation. *B. longum*-mediated VRE inhibition is significantly augmented by lactulose, potentially explaining the expansion of VRE seen in fecal samples lacking expanded *Bifidobacteria* populations despite lactulose administration. Consistent with VRE expansion seen in patients treated with lactulose without *Bifidobacteria* expansion, VRE growth is also augmented by lactulose (Fig S7E,F).

### Bifidobacteria expansion and associated metabolite production correlates with reduced incidence of infections in advanced liver disease

To determine whether lactulose-mediated *Bifidobacteria* expansion and associated metabolite changes are associated with clinical benefits, we correlated the gut microbiome metagenomic and metabolic data with development of common infections in cirrhosis, including spontaneous bacterial peritonitis (SBP) and bacteremia (Figs 6, S8, and S9).

**Figure 6:**
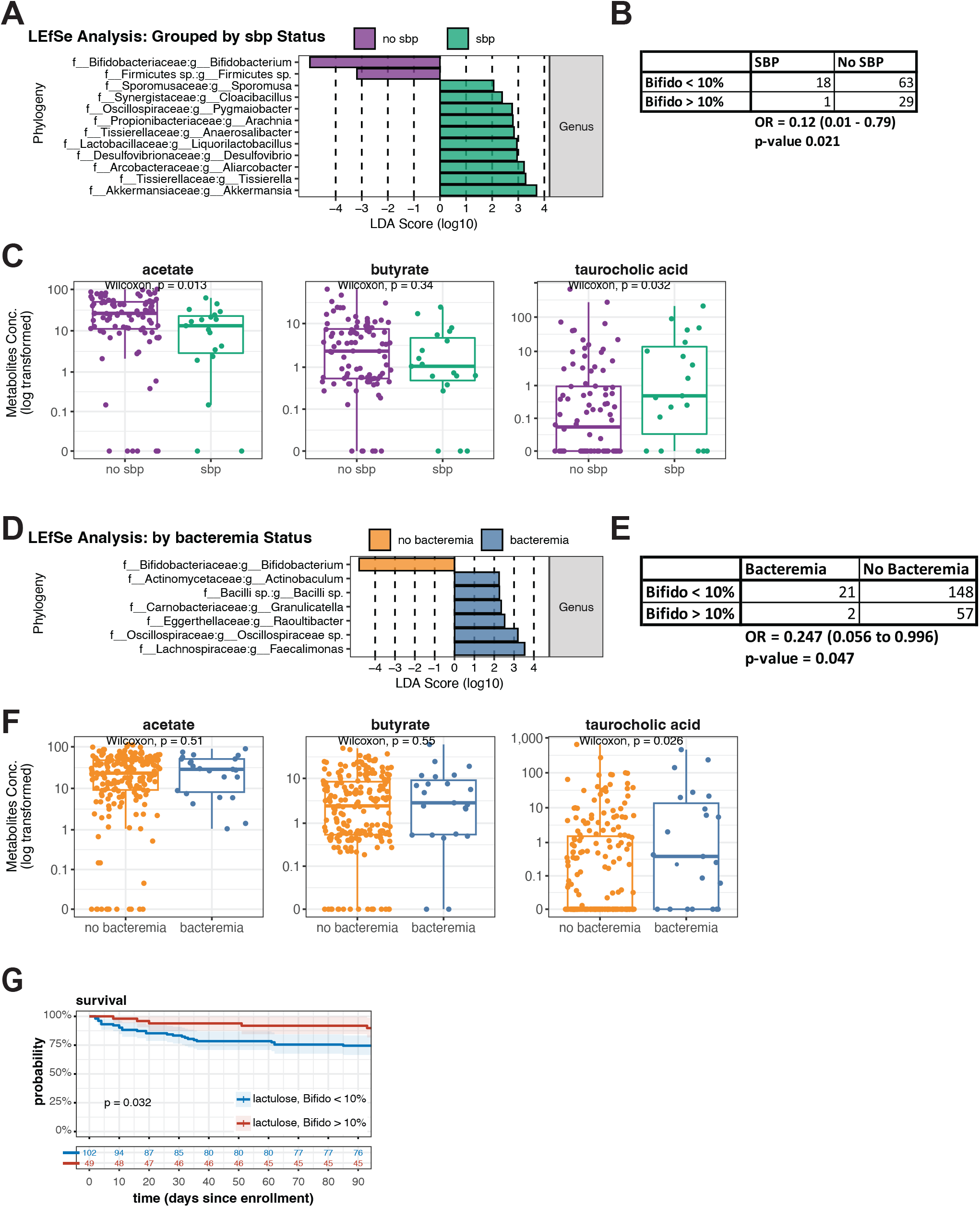
*Bifidobacteria* expansion and associated metabolite production are associated with decreased incidence of spontaneous bacterial peritonitis (SBP) and bacteremia and prolonged survival. (A) A total of 19 episodes of SBP were identified that had an adjacent stool sample. A Linear discriminant analysis effect size (LEfSe) showing the significant (Wilcoxon rank-sum, two-tailed, p ≤ 0.05) effect sizes of taxa between groups (purple bars: no SBP, green bars: SBP). (B) Odds ratio of being diagnosed with SBP if *Bifidobacteria* relative abundance is greater than 10% was calculated from the data shown. (C) When compared to fecal samples from patients with SBP, those without SBP had statistically (Wilcoxon rank-sum, two-tailed, p < 0.05) higher concentrations of acetate, and lower concentrations of the conjugated primary bile acid taurocholic acid. (D) A total of 23 episodes of bacteremia were identified from 228 unique blood cultures that had a stool sample within the pre-defined time window relative to blood culture. A Linear discriminant analysis effect size (LEfSe) showing the significant (Wilcoxon rank-sum, two-tailed, p ≤ 0.05) effect sizes of taxa between groups (yellow bars: no bacteremia, blue bars: bacteremia). (E) Odds ratio of being diagnosed with bacteremia if *Bifidobacteria* relative abundance is greater than 10% was calculated from the data shown. (F) When compared to fecal samples from patients with bacteremia, those without bacteremia had statistically (Wilcoxon rank-sum, two-tailed, p < 0.05) similar levels of short chain fatty acids and lower concentrations of the conjugated primary bile acid taurocholic acid. Units for SCFA are mM, and units for BA derivatives are in μg/mL. (G) Kaplan-Meier curves were generated for survival based on initial stool sample characteristics. Survival curves were stratified based on lactulose administration and *Bifidobacteria* expansion of the initial stool sample. The number at risk at each time-point is shown below.

We identified 111 ascites samples with near concurrent fecal samples for analysis, among which there were 19 diagnoses of SBP (Fig 6A, B, S8A, Table S1). LEfSe analysis demonstrated that *Bifidobacteria* was strongly associated with resistance to SBP, while presence of the mucus-degrading *Akkermansia* was associated with SBP (Fig 6A). We used a frequency cutoff of 10% to distinguish patients with high *Bifidobacteria* from those with low *Bifidobacteria* frequencies. We found that 1 of 29 patients (3.4%) with ≥10% Bifidobacteria frequency was diagnosed with SBP while 18 of 63 patients (28.6%) with less than 10% Bifidobacteria developed SBP (Fig 6B; Odds Ratio 0.12, 95% CI 0.01 – 0.79, p value 0.021). Qualitative metabolomics demonstrated that glycholithocholic acid and serotonin were significantly associated with being SBP-free (Fig S8B). The *Bifidobacteria-*generated SCFA acetate was also relatively increased in SBP-free samples, albeit to a lesser extent (p < 0.05, log_2_ fold-change 0.9, Fig S8B). Quantitative metabolomics demonstrated that acetate was significantly higher in SBP-free fecal samples and the conjugated primary bile acid taurocholic acid was significantly higher in samples with SBP (Fig 6C). Consistent with a role for *Bifidobacteria* in protection from SBP, samples from patients without SBP had a significantly greater conversion of conjugated to unconjugated primary BA (CA:GCA ratio) but similar conversion of primary to secondary BA (Fig S8D).

To correlate fecal microbiome compositions and metabolite concentrations with development of bacteremia, we paired 328 blood cultures with fecal samples for analysis. Blood culturing identified 23 distinct diagnoses of bacteremia (Fig 6D, E, S9A, Table S2) and LEfSe analysis demonstrated that *Bifidobacteria* was associated with remaining bacteremia-free (Fig 6D). There were 2 diagnoses of bacteremia in 57 patients (3.5%) with ≥10% *Bifidobacteria* and 21 of 148 patients (14.2%) with <10% *Bifidobacteria* (Fig 6E; Odds Ratio 0.247, 95% CI 0.056 – 0.996, p value 0.047). Qualitative metabolomics demonstrated that conjugated primary BA (glycocholic, taurocholic, and glycochenodeoxycholic acid) and the primary BA cholic acid were all significantly increased in bacteremia-associated samples (Fig S9B). Targeted quantitative metabolomics confirmed that samples from bacteremic patients had reduced capacity to hydrolyze conjugated bile acids (Fig 6F, S9D). These data suggest that lactulose-mediated *Bifidobacteria* expansion with resultant SCFA production and bile salt hydrolase activity may play a protective role against some of the most common and devastating infectious complications of advanced liver disease.

### Lactulose-mediated gut Bifidobacteria expansion is associated with improved transplant-free survival

Finally, we determined whether initial stool characteristics can identify patients at increased risk of adverse outcomes and thus might serve as targets for microbiome-based therapies. To do so, we assessed 90-day overall survival in patients stratified by lactulose exposure and *Bifidobacteria* expansion. Compared to patients who received lactulose but did not expand *Bifidobacteria*, patients with lactulose-associated *Bifidobacteria* expansion had significantly improved overall 90-day survival (Fig 6G) with similar rates of liver transplantation (Fig S10). While many patient characteristics between the groups were similar (including age, race, BMI, cause of liver disease, and presence of clinically significant portal hypertension), patients who received lactulose but did not expand *Bifidobacteria* had clinical parameters that correlate with a poor prognosis (including higher MELD-Na scores and higher incidence of additional organ failures present upon hospitalization) (Table 3). This suggests that *Bifidobacteria* expansion and associated metabolite profiles can serve as a biomarker for healthier patient cohort, which may be due to the gut microbiome differences along with other disease-specific factors.

## DISCUSSION

Lactulose has been used to treat hepatic encephalopathy for over 50 years^66^. Despite widespread use and guideline recommendations for HE treatment from medical societies, its mechanism of action is incompletely understood^14,67^. While lactulose is postulated to provide benefits by decreasing bowel transit time and gut lumen acidification, thereby decreasing ammonia absorption, its role in altering the microbiome has remained largely undefined. Herein, we demonstrate that in patients with liver disease, lactulose administration, in the absence of broad-spectrum antibiotic administration, leads to expansion of *Bifidobacteria* species. This expansion results in a distinct gut microbiome taxonomic and metabolic profile and is associated with exclusion of antibiotic-resistant pathobionts. *Bifidobacteria* inhibit growth of antibiotic-resistant Enterococcal and Enterobacterales species, effects that are augmented by lactulose. Our findings suggest a protective role of lactulose-mediated *Bifidobacteria* expansion in patients with liver disease. Consistent with this, *Bifidobacteria* expansion and associated metabolites are associated with reduced incidence of SBP and bacteremia as well as improved 90-day survival.

In addition to treating precipitating factors, such as systemic infection, lactulose has largely replaced dietary modification, antibiotics, and laxatives as the first line treatment for hepatic encephalopathy. All HE therapies are aimed at reducing gut ammonia production and absorption, and the initial reports of lactulose use for HE concluded that this is due to luminal acidification^66,68^. It has also been assumed that, similar to healthy subjects and experimental models ^12,69,70^, lactulose exhibits a prebiotic effect, but this has not been shown in patients with liver disease^7,10,66,71^. The authors of the initial trial of lactulose were unable to detect changes in fecal bacteria^66^; however, they were limited by culture-based techniques. More recent studies characterizing the microbiomes of cirrhotic patients did not reveal Bifidobacteria expansion in patients treated with lactulose ^9^, most likely because 16S rRNA sequencing platforms often do not amplify Bifidobacterium 16S rRNA genes^72^. Along similar lines, lactulose withdrawal demonstrated only slight decreases in *Faecalibacterium* species using sequencing techniques^10^. Our study avoided 16S rRNA amplification bias by metagenomically sequencing fecal samples and demonstrates marked expansion of *Bifidobacteria* in a large subset of cirrhotic patients treated with lactulose.

*Bifidobacterium* species are dominant fecal microbes in breast fed infants and are commonly thought of as prototypical health-promoting bacteria^73^. *Bifidobacterium* species imprint the human immune system^74^, are associated with decreased development of atopy and autoimmune diseases^75^, and administration decreases rates of necrotizing enterocolitis (NEC) in preterm infants^76^. In adults, fecal *Bifidobacteria* are thought to modulate anti-tumor immunity and enhance immunotherapy efficacy in humans with melanoma and synergize with immunotherapies to reduce melanoma growth in mouse models^77,78^. *Bifidobacteria* also produce acetate and lactic acid, which antagonize pathogens to reduce the incidence of enteric infections in infants. While *Bifidobacteria* does not provide colonization resistance to pathogenic *E. coli* in a murine model, specific acetate-producing *Bifidobacteria* strains limit systemic disease by improving mucosal integrity via acetate promotion of epithelial integrity and from heightened immune surveillance^64^.Gut acidification, mucosal barrier enhancement, and immunomodulatory effects of *Bifidobacteria* may all benefit patients with liver disease, who are immunocompromised and at high risk of enteric pathogen colonization, growth and dissemination.

While *Bifidobacteria* attenuate liver disease progression in rodent models^79,80^, our study suggests that *Bifidobacteria* may reduce the complications of severe liver disease and thus may enhance approaches for supportive care. We report an association between *Bifidobacteria* expansion and reduced incidence of systemic infection and prolonged patient survival. This beneficial association has multiple plausible explanations that may be driven by the unique metabolite profile including increased acetate levels and enhanced hydrolysis of conjugated primary bile acids. Similar metabolite profiles appear to be protective against both SBP and bacteremia. Short chain fatty acid production has been linked to gut colonization resistance of common gram-negative enteric pathogens^81^. Similarly, *Bifidobacteria* expansion was associated with reduced abundance of the common gram-positive pathogen, VRE in our patients, and inhibited VRE growth in vitro, likely via both media acidification and SCFA production. *Bifidobacteria*-mediated VRE growth inhibition was significantly enhanced by lactulose, which also significantly increased SCFA production and acidification in vitro and acetate production in both mice and patients. While colonization and expansion are important initial steps in infection development, SCFAs are also known to promote intestinal barrier function by enhancing both epithelial integrity and mucosal immune function, thereby limiting pathogen translocation ^28,29,64,82^. Moreover, *Bifidobacteria* are associated with decreased intestinal permeability in patients with alcohol dependence, which is extremely common in our study population^83^. In our study, *Bifidobacteria*-expanded samples also had significantly higher levels of indole-3-carboxaldehyde, which activates the aryl hydrocarbon receptor to increase mucosal IL-22 production and maintain mucosal reactivity towards pathogens.

Hospitalized patients with liver disease are commonly treated with broad spectrum antibiotics with accompanying marked microbiome derangements, and our findings will aid in identifying patients who may benefit from targeted microbiome therapy. Success of a biotherapeutic approach to supportive care likely depends on both the abundance of *Bifidobacteria* in the patient’s gut at the time of treatment initiation and co-administration of a prebiotic, such as lactulose^65^. In subjects without liver disease, both prebiotic and probiotic approaches result in *Bifidobacteria* expansion, and a synbiotic approach prolongs functional *Bifidobacteria* engraftment^12,84,85^. In outpatients with liver disease, synbiotic approaches with multiple lactic acid bacteria and fermentable fiber sources reduce the blood ammonia levels, endotoxinemia, and the incidence of hepatic encephalopathy^86,87^. These studies do not assess the durability or metabolic function of the administered probiotic or correlate use with other outcomes in liver disease. Another important consideration that has not been well-studied even in healthy adults is the functional (i.e. metabolomic) change that marked *Bifidobacteria* expansion has on the gut microbiome.

While associated with improved outcomes in our population, a *Bifidobacteria*-expanded microbiome is not reflective of a normal, healthy adult microbiome with regards to microbial composition or metabolite profiles. While acetate has been shown to prevent gut pathogen dissemination in a murine model, butyrate is better studied with regard to epithelial barrier function and mucosal immune function^28,29,31,64^. Secondary bile acids are not only often considered markers of a “healthy” gut microbiome but have also been implicated in prevention of enteric infections^40^ and reducing intestinal inflammation^88^. This will also be an important consideration in rational probiotic therapy design, and supplementation with multiple bacterial species with complementary metabolite production is likely the preferred approach. Conversely, *Bifidobacteria* are an important component of the developing human gut microbiome. Lactulose-mediated *Bifidobacteria* expansion may therefore be a stepping-stone to allow for expansion and engraftment of additional beneficial commensal organisms and restoration of a microbiome more similar to healthy adults. A recent paper studying the effect of active alcohol consumption on the murine gut microbiome concluded that acetate produced during alcohol metabolism results in increased *Bacteroidetes*^89^, and it is plausible to think that *Bifidobacteria*-derived metabolites may also allow for commensal expansion.

In conclusion, we report here that lactulose-mediated gut *Bifidobacteria* expansion is associated with a unique fecal metagenomic and metabolic profile, reduced clinically-relevant antibiotic-resistant pathobiont burden, and improved clinical outcomes in hospitalized patients with liver disease, including reduced incidence of SBP and bacteremia and prolonged survival. Our study provides novel insights into the mechanism of action of a liberally prescribed medication and rationale for future studies designed to precisely target the gut microbiome to prevent complications of advanced liver disease.

## Data Availability

In addition to the repositories specified below, all raw data included in the manuscript is publicly available at: https://github.com/yingeddi2008/DFILiverDiseaseMicrobiome.
1. Metagenomic information is publicly available on NCBI under BioProject ID PRJNA912122.
2. Quantitative fecal metabolomic information paired to fecal metagenomic information is publicly available on NCBI under BioProject ID PRJNA912122. 
3. Raw data files are publicly available on MetaboLights project ID MTBLS7046.

## GRAPHICAL ABSTRACT

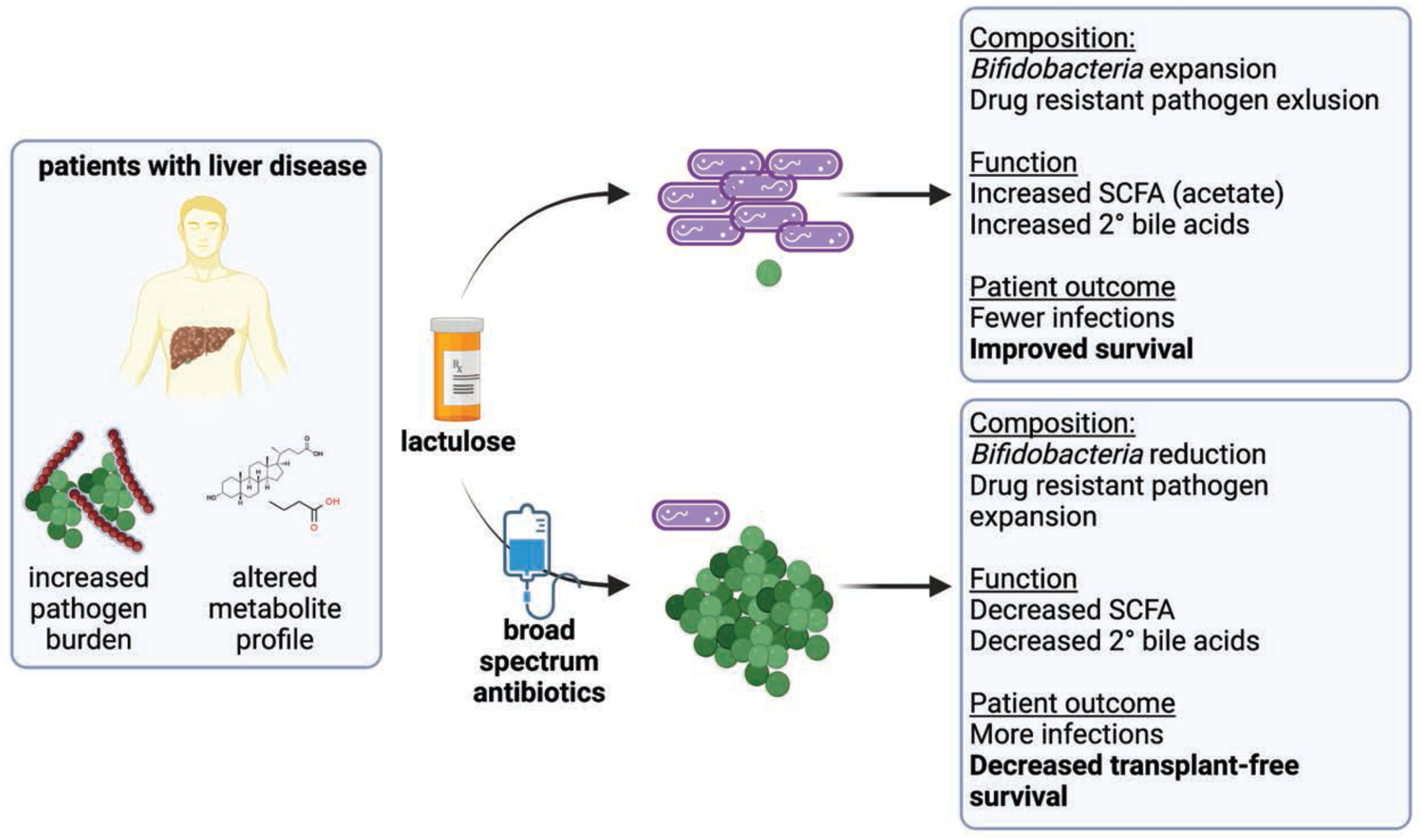

---

| **n** | **263** |
| --- | --- |
| Age (median [IQR]) | 58.40 [46.75, 66.45] |
| Sex = Male (%) | 154 (58.6) |
| Race (%) |  |
| African American | 86 (32.7) |
| Asian / Pacific Islander | 12 ( 4.6) |
| Caucasian | 126 (47.9) |
| Hispanic | 34 (12.9) |
| Other | 5 ( 1.9) |
| BMI (median [IQR]) | 27.77 [23.53, 33.10] |
| Liver Disease Etiology (%) |  |
| Alcohol | 134 (51.0) |
| Vascular | 30 (11.4) |
| NASH | 27 (10.3) |
| HCV | 16 ( 6.1) |
| Autoimmune | 12 ( 4.6) |
| Biliary | 8 ( 3.0) |
| DILI | 6 ( 2.3) |
| CPI Hepatitis | 5 ( 1.9) |
| Metabolic | 4 ( 1.5) |
| HBV | 3 ( 1.1) |
| Other | 18 ( 6.8) |
| MELD-Na (median [IQR]) | 18.69 [10.61, 27.88] |
| Clinically significant portal HTN (%) | 183 (69.6) |
| HCC (%) | 27 (10.3) |
| End organ dysfunction (%) | 137 (52.1) |
| Hepatic Encephalopathy (%) | 59 (22.4) |
| Circulatory Dysfunction (%) | 32 (12.2) |
| Respiratory Dysfunction (%) | 38 (14.4) |
| Renal Dysfunction (%) | 85 (32.3) |

|  | lactulose, Bifido < 10% | lactulose, Bifido > 10% | p | test |
| --- | --- | --- | --- | --- |
| n | 102 | 49 |  |  |
| Age (median [IQR]) | 57.80 [49.45, 64.68] | 62.50 [51.30, 67.30] | 0.196 | nonnorm |
| Sex = Male (%) | 67 (65.7) | 24 ( 49.0) | 0.074 |  |
| Race (%) |  |  | 0.556 |  |
| African American | 30 (29.4) | 16 ( 32.7) |  |  |
| Asian / Pacific Islander | 3 ( 2.9) | 4 ( 8.2) |  |  |
| Caucasian | 57 (55.9) | 22 ( 44.9) |  |  |
| Hispanic | 10 ( 9.8) | 6 ( 12.2) |  |  |
| Other | 2 ( 2.0) | 1 ( 2.0) |  |  |
| BMI (median [IQR]) | 28.60 [24.13, 34.01] | 28.72 [23.25, 33.27] | 0.534 | nonnorm |
| Liver Disease Etiology (%) |  |  | 0.163 |  |
| Alcohol | 68 (66.7) | 25 ( 51.0) |  |  |
| NASH | 10 ( 9.8) | 10 ( 20.4) |  |  |
| HCV | 4 ( 3.9) | 5 ( 10.2) |  |  |
| Vascular | 6 ( 5.9) | 2 ( 4.1) |  |  |
| DILI | 4 ( 3.9) | 0 ( 0.0) |  |  |
| Autoimmune | 1 ( 1.0) | 1 ( 2.0) |  |  |
| CPI Hepatitis | 1 ( 1.0) | 0 ( 0.0) |  |  |
| HBV | 1 ( 1.0) | 1 ( 2.0) |  |  |
| Metabolic | 2 ( 2.0) | 0 ( 0.0) |  |  |
| Biliary | 2 ( 2.0) | 1 ( 2.0) |  |  |
| other | 3 ( 2.9) | 4 ( 8.2) |  |  |
| MELD-Na (median [IQR]) | 25.50 [16.18, 35.38] | 18.85 [12.26, 25.75] | 0.002 | nonnorm |
| Clinically significant portal HTN (%) | 88 (86.3) | 43 ( 87.8) | 1 |  |
| HCC (%) | 3 (2.9) | 9 ( 18.4) | 0.003 |  |
| End organ dysfunction (%) | 81 (79.4) | 21 (42.9) | <0.001 |  |
| Hepatic Encephalopathy (%) | 45 (44.1) | 11 (22.4) | 0.016 |  |
| Circulatory Dysfunction (%) | 19 (18.6) | 1 (2.0) | 0.011 |  |
| Respiratory Dysfunction (%) | 23 (22.5) | 4 (8.2) | 0.053 |  |
| Renal Dysfunction (%) | 55 (53.9) | 9 (18.4) | <0.001 |  |
| lactulose, Bifido > 10% (%) | 0 ( 0.0) | 49 (100.0) | <0.001 |  |

## SUPPLEMENTAL FIGURE LEGENDS

**Figure S1:**
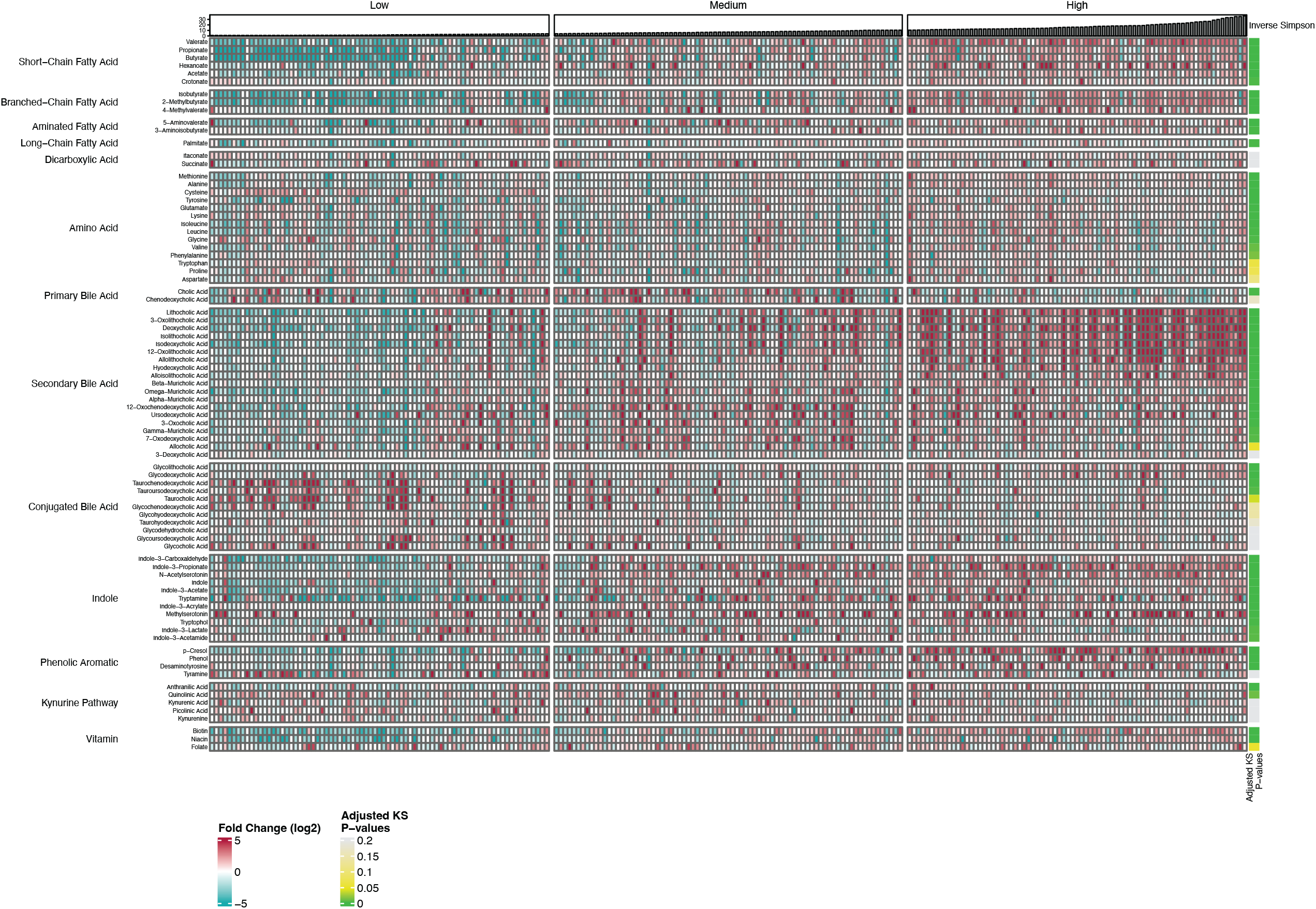
Fecal samples from patients with liver disease display a wide range of metabolomic profiles that correlates with metagenomic alpha-diversity. Initial samples from each patient were arranged in order of increasing alpha-diversity as measured by inverse Simpson from left-to-right and shown in the top panel. One third of samples were grouped into low, medium, or high alpha-diversity. Eighty-two metabolites were analyzed qualitatively for each sample, and the relative concentrations are represented in the pseudocolored heat map. For each compound, a Kruskal-Wallis test was run between the low, medium, and high diversity groups, and the statistics are color-coded in the far-right column. Samples with high alpha-diversity tended to have increased levels of SCFAs, secondary bile acids, and select indole derivatives compared to samples with lower alpha-diversity.

**Figure S2:**
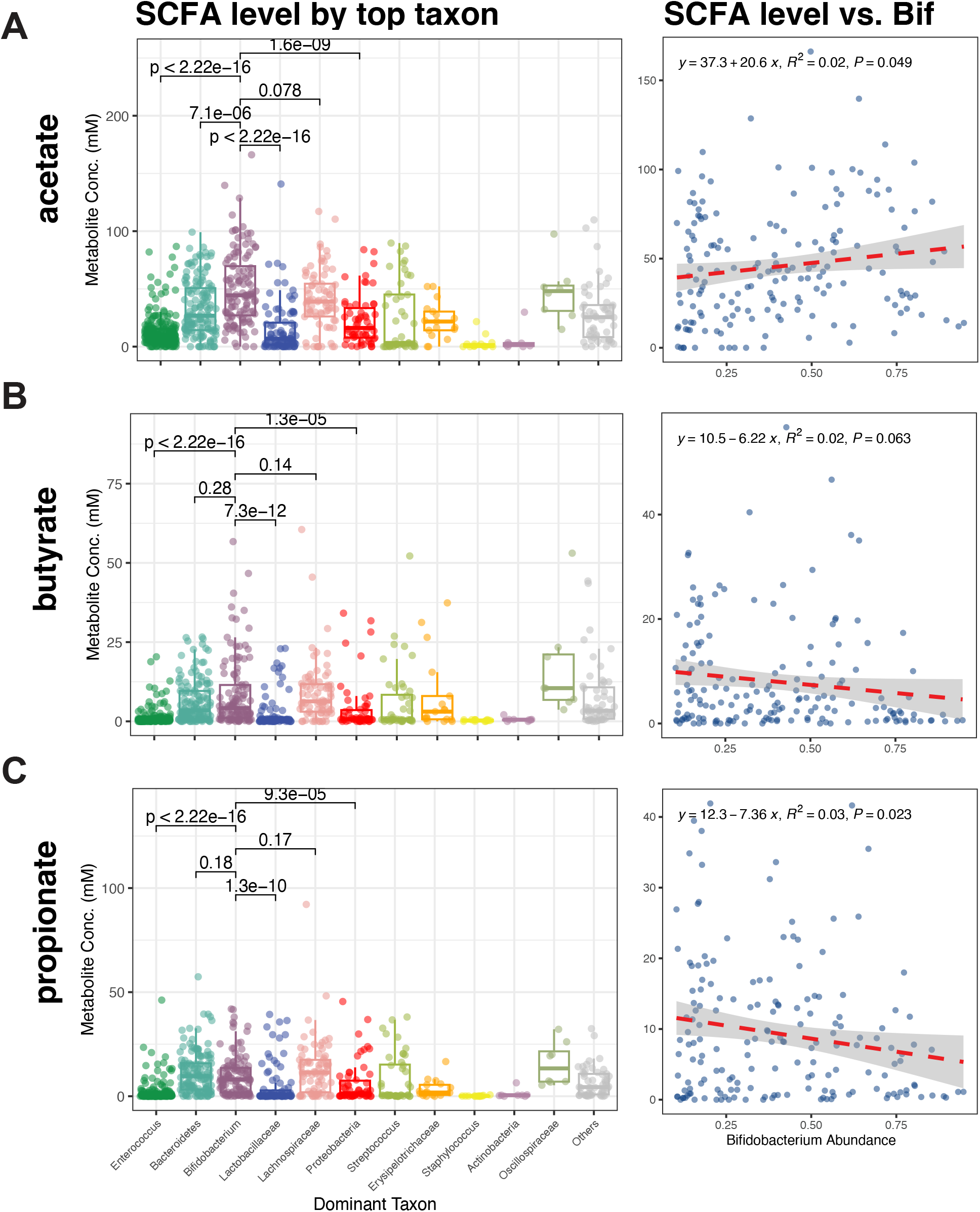
*Bifidobacteria* expansion is associated with increased acetate production. (A-C) Graphs (left column) show the indicated SCFA levels for each sample grouped by the most abundant taxon as shown in taxonomic UMAP in Figure 2A. In the bar graphs, each individual point represents a single stool sample. Median and interquartile range are indicated by the line and box, respectively. Global statistics were done using the Kruskal–Wallis test and statistical comparisons between individual groups were analyzed using the Wilcoxin rank sum. Individual groups were compared to the *Bifidobacteria* dominated group, the unique cluster in this patient cohort. In the right-most graphs, the indicated short chain fatty acid is plotted as a function of *Bifidobacteria* abundance in samples containing ≥ 10% *Bifidobacteria*. Linear regression with 95% confidence interval is plotted.

**Figure S3:**
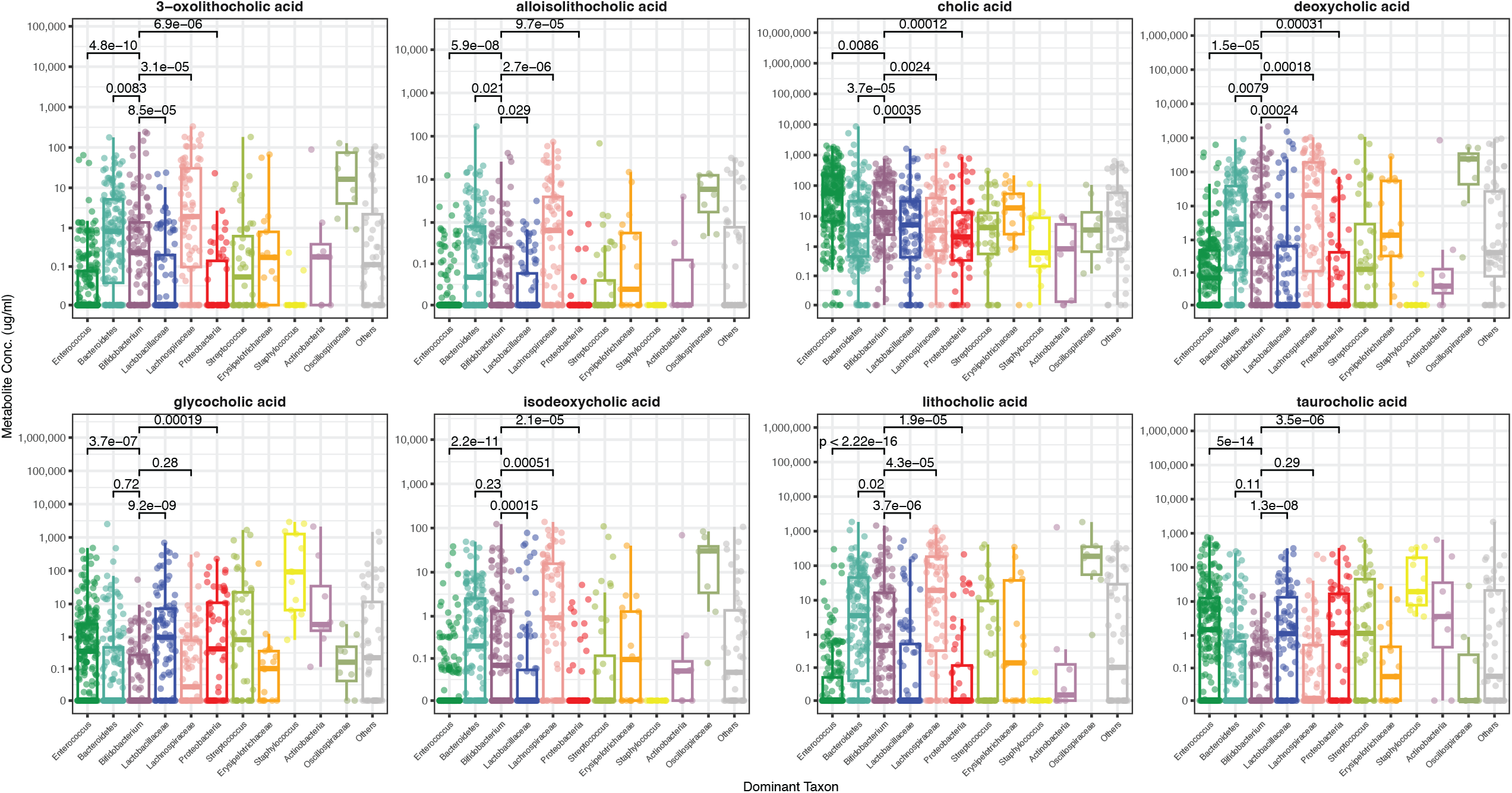
*Bifidobacteria* expansion correlates with increased capacity to deconjugate bile acids. Graphs show the BA levels for each sample grouped by the most abundant taxon as shown in the taxonomic UMAP in Figure 2A. In the bar graphs, each individual point represents a single stool sample. Median and interquartile range are indicated by the line and box, respectively. Global statistics were done using the Kruskal–Wallis test and statistical comparisons between individual groups were analyzed using the Wilcoxin rank sum. Individual groups were compared to the *Bifidobacteria* dominated group, the unique cluster in this patient cohort.

**Figure S4:**
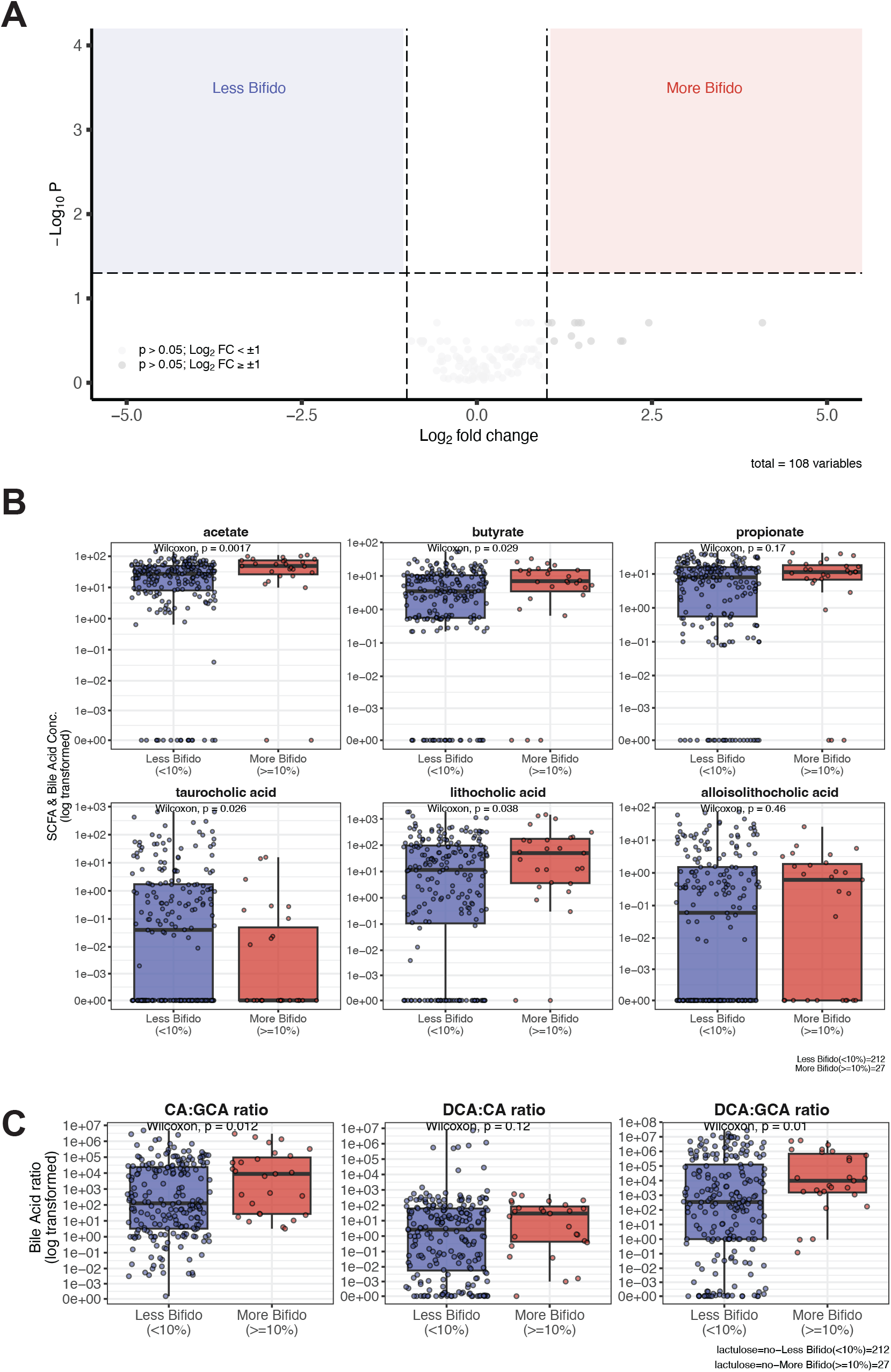
In the absence of lactulose, *Bifidobacteria* expansion is associated with modest fecal metabolite changes. (A) Volcano plot (log_2_fold change vs. log_10_p-value) of qualitative metabolites comparing samples with low (< 10%) vs. high (≥10%) *Bifidobacteria* abundance after lactulose exposure. P-values are corrected for multiple comparisons. Values with log2 fold-change > 1 (corresponding to a 2-fold change with a p-value < 0.05 were considered significant. No values were significantly different between the two groups without lactulose exposure. (B) Bar graphs for select SCFA (acetate, butyrate, and propionate) and bile acids (taurocholic, lithocholic, and alloisolithocholic acid) where each point represents a single value. Median and interquartile range are indicated by the line and box, respectively. Statistical comparisons between groups were analyzed using the Wilcoxin rank sum. Units for SCFA are mM, and units for BA derivatives are in Dg/mL. (C) Bile acid conversion from conjugated-primary BA to primary BA and then to secondary BAs was tested for each sample. Each individual point represents a ratio for an individual sample. Samples were grouped by whether they had expanded *Bifidobacteria* in the absence of lactulose. Statistical comparisons between groups were analyzed using the Wilcoxin rank sum. CA: cholic acid; GCA: glycocholic acid; DCA: deoxycholic acid. No ratio was significantly different.

**Figure S5:**
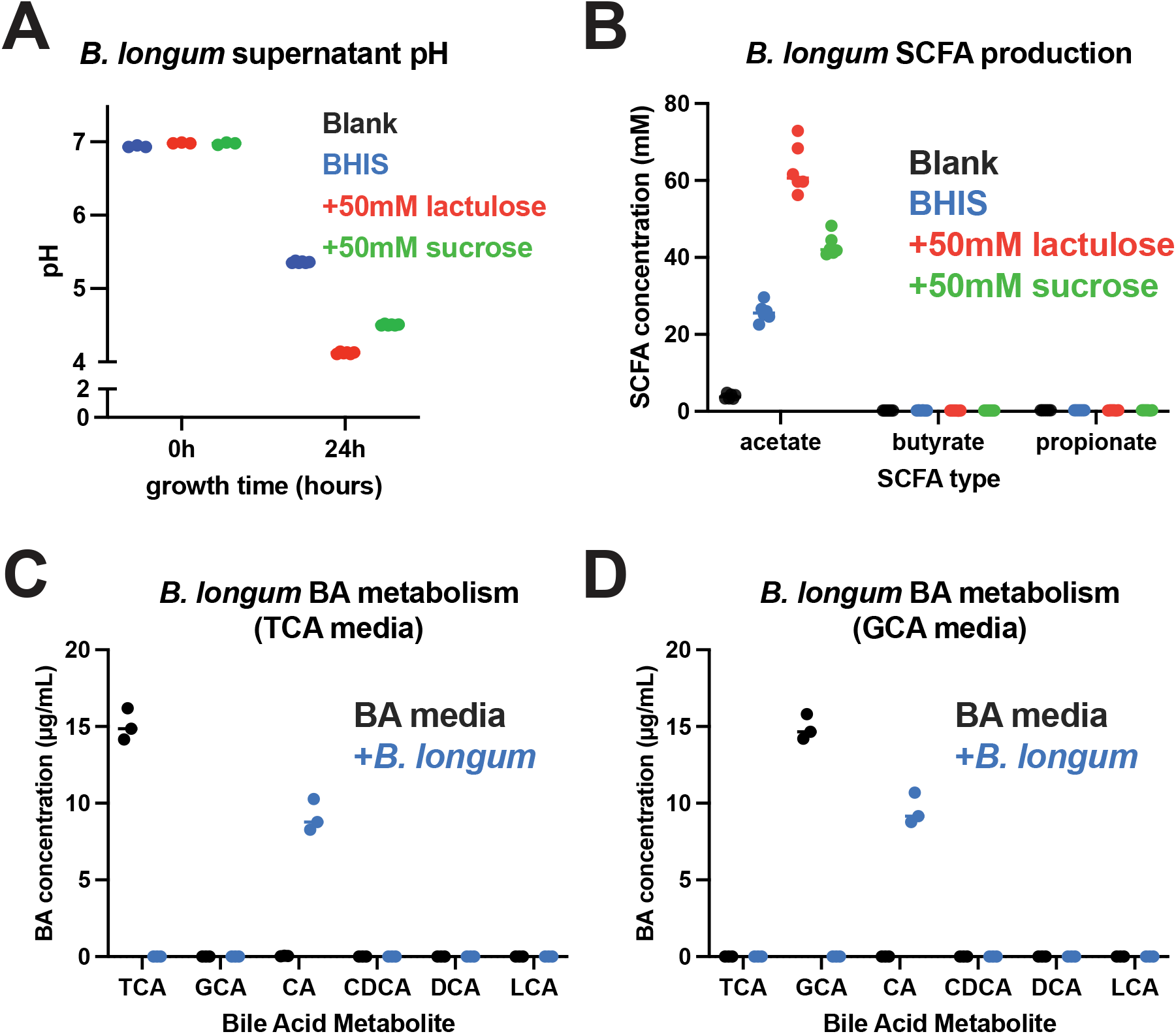
*Bifidobacterium longum* acidifies pH, produces acetate, and efficiently hydrolyzes conjugated primary bile acids *in vitro*. *B. longum* was grown in regular BHIS media (blue) or BHIS supplemented with 50mM lactulose (red) or 50mM sucrose (green). Before and after 24 hours of growth in each BHIS media condition, (A) pH was measured. (B) SCFA concentrations were measured after 24 hours of growth in the indicated media. (C and D) *B. longum* was grown for 24 hours in BHIS supplemented with 10μg/ml of conjugated primary bile acid (either (C) taurocholic or (D) glycocholic acid), and targeted, quantitative bile acids were measured in the supernatant. All plots are representative of three independent experiments done in at least triplicate. TCA: taurocholic acid, GCA: glycocholic acid, CA: cholic acid, CDCA: chenodeoxycholic acid, DCA: deoxycholic acid, and LCA: lithocholic acid.

**Figure S6:**
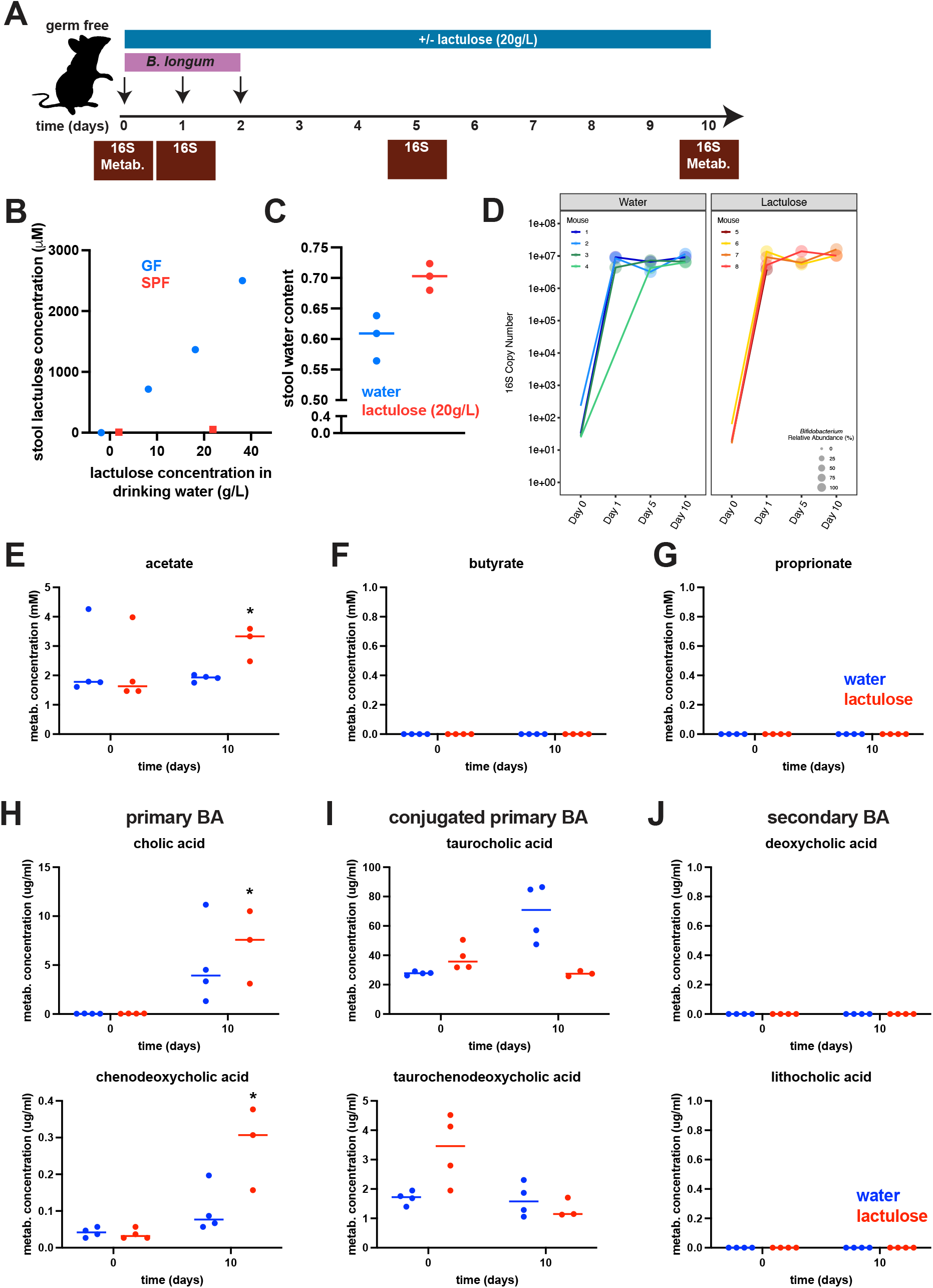
*B. longum* colonizes germ free mice and displays similar metabolic activity *in vivo*. (A) Schematic indicating timing of *B. longum* gavage, lactulose administration, and stool collection in ex-germ free mice. 16S: metagenomics; Metab.: targeted, quantitative metabolomics. (B) Stool lactulose was measured in germ free mice receiving regular drinking water or water containing 10, 20, or 40g/L or in SPF mice receiving either regular water or 20g/L of drinking water. (C) Stool water concentration of GF mice receiving regular drinking water or water containing 20g/L of lactulose was measured. (D) Quantitative 16S metagenomics over time of stool samples from ex-GF mice colonized with *B. longum* who were fed either regular water or water containing 20g/L lactulose. There was no statistical difference in *Bifidobacteria* 16S counts between water and lactulose-fed mice at each time point. (E – G) Short chain fatty acid concentrations were measured before and 10-days after *Bifidobacteria* inoculation in each mouse. *, p < 0.05 by t-test comparing water to lactulose-treated mice at a given time point. (H) Primary, (I) conjugated primary, and (J) secondary bile acids were measured at indicated timepoints in each mouse. Graphs depict each stool sample as an individual dot, and the line indicates the mean +/-standard error of the mean. *, p < 0.05 by t-test comparing to time 0 for a given treatment (lactulose or water).

**Figure S7:**
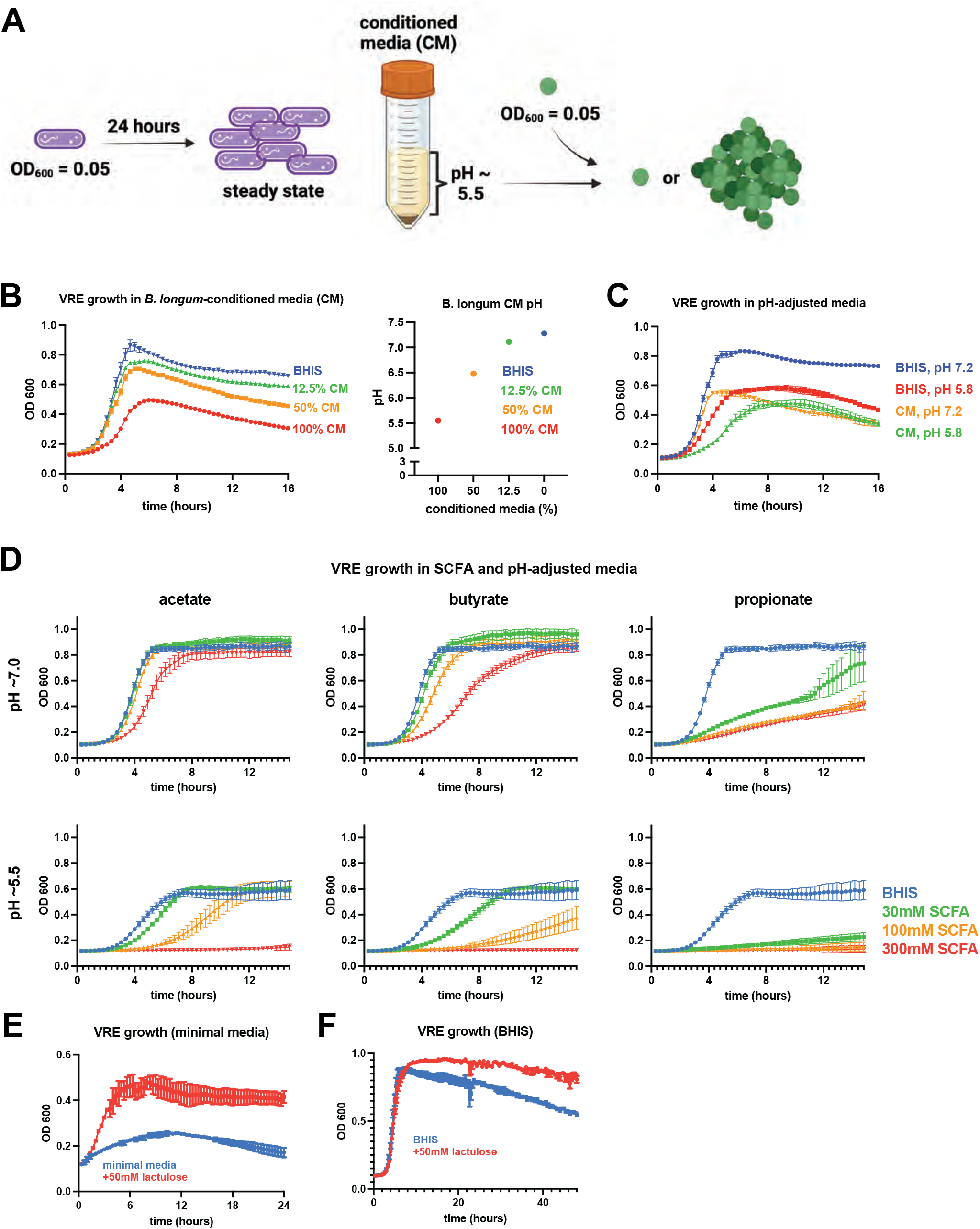
*Bifidobacterium longum* supernatant inhibits VRE growth *in vitro*. (A) Schematic of experimental design for VRE growth in *B. longum* conditioned media (CM). *B. longum* was grown in BHIS for 24-hours prior to collecting and filtering the supernatant. VRE growing at steady state was then diluted to a low density (OD_600_ = 0.05) prior to inoculating in various dilutions of *B. longum* conditioned media in fresh BHIS. (B) VRE growth curves in 100%, 50%, 12.5%, and 0% CM with the pH of the 4 concentrations of CM in BHIS shown in the right portion of this panel. (C) The pH of neutral BHIS was acidified from 7.2 to 5.8 using HCl, and the pH of *B. longum* CM was neutralized from 5.8 to 7.2 with NaOH prior to VRE inoculation. Growth curves for VRE in each of these 4 conditions are shown. (D) VRE was inoculated into either neutral (top panels) or acidified (bottom panels) BHIS containing 0mM (blue), 30mM (green), 100mM (orange), or 300mM (red) of the indicated SCFA. (E) VRE was grown in minimal media containing either no additive (blue) or 50mM lactulose, and OD_600_ was measured over time. (F) VRE was grown in BHIS with or without 50mM lactulose, and OD_600_ was measured over time. All plots are representative of three independent experiments done in at least triplicate.

**Figure S8:**
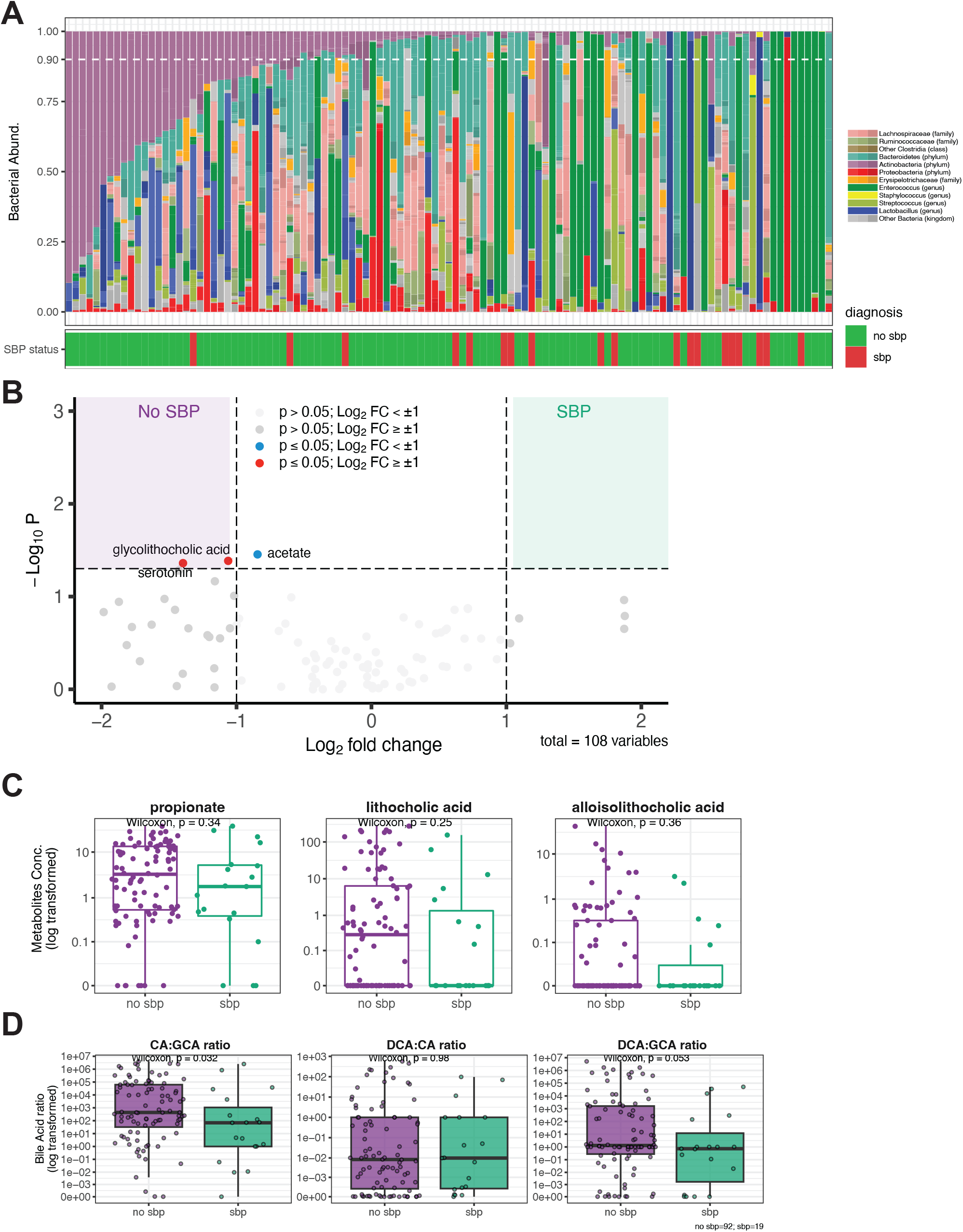
*Bifidobacteria* expansion and associated metabolite production are associated with decreased incidence of spontaneous bacterial peritonitis (SBP). (A) Taxonomic relative abundance by shotgun metagenomics of stool samples paired to ascites samples is plotted in order of decreasing *Bifidobacteria* abundance from left to right. Underneath each stool sample is the clinical diagnosis associated with an ascites sample (SBP (red) or not SBP (green)). (B) Volcano plot of normalized metabolite concentrations. Significant values had log_2_ fold-change values ≥ 1 and p-value < 0.05. (C) Quantitative levels for propionate, lithocholic acid, and alloisolithocholic acid were compared from samples associated and not associated with SBP. Units for SCFA are mM, and units for BA derivatives are in μg/mL. (D) Ratios of primary-conjugated (glycocholic acid, GA), primary (cholic acid, CA), and secondary BAs (deoxycholic acid, DCA) were generated for each sample, and each point represents a single sample. Box plots depict median and IQR, and statistics were performed using a Wilcoxon rank-sum, two-tailed, adjusted p-value.

**Figure S9:**
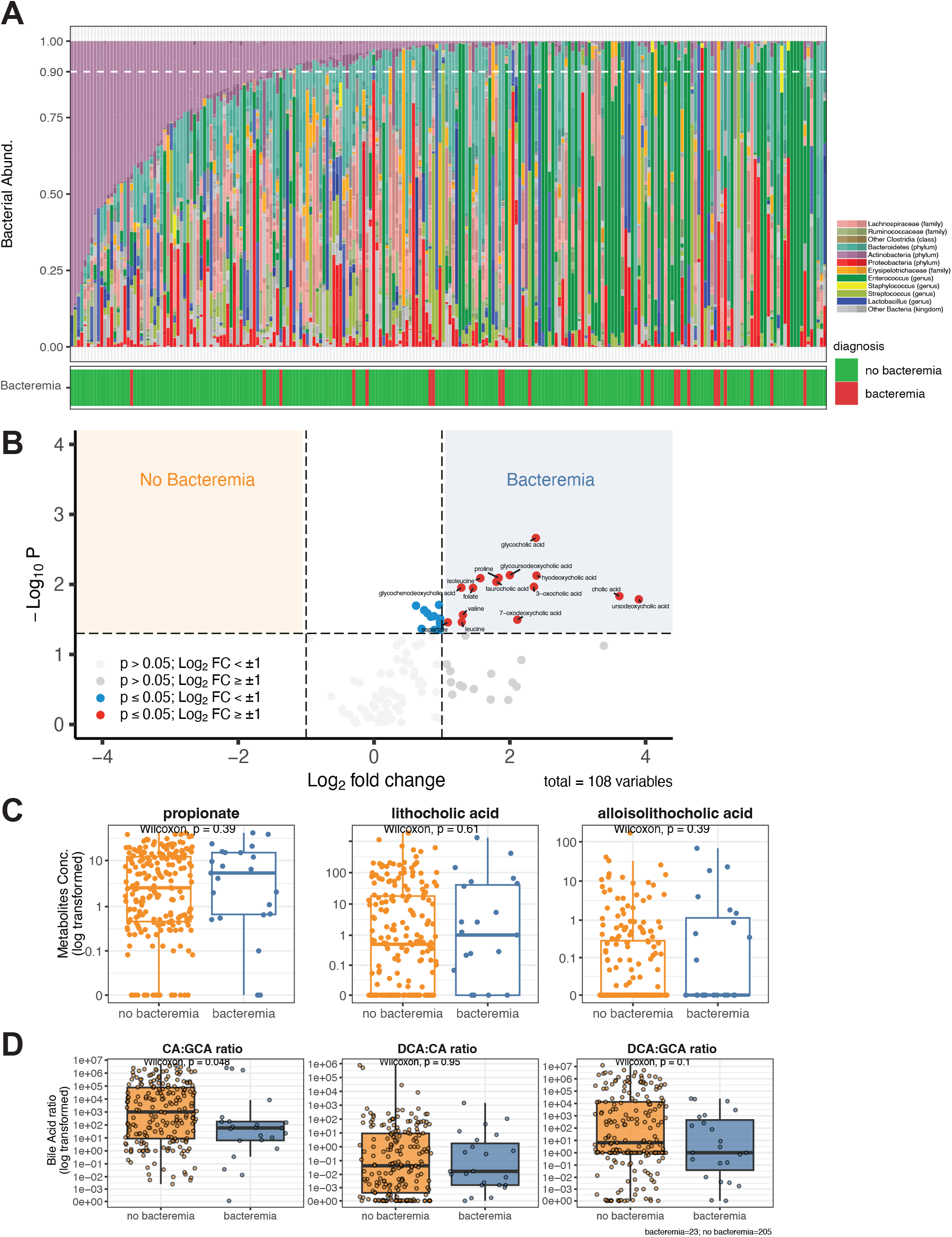
*Bifidobacteria* expansion and associated metabolite production are associated with decreased incidence of bacteremia. (A) Taxonomic relative abundance by shotgun metagenomics of stool samples paired to blood cultures is plotted in order of decreasing *Bifidobacteria* abundance from left to right. Underneath each stool sample is the clinical diagnosis associated with a given blood culture (bacteremia (red) or no bacteremia (green)). (B) Volcano plot of normalized metabolite concentrations. Significant values had log_2_ fold-change values ≥ 1 and p-value < 0.05. These included multiple conjugated primary bile acids (e.g. glycocholic acid and taurocholic acid). (C) Quantitative metabolites for propionate, lithocholic acid, and alloisolithocholic acid were compared from samples associated and not associated with bacteremia. Units for SCFA are mM, and units for BA derivatives are in μg/mL. (D) Ratios of primary-conjugated (glycocholic acid, GA), primary (cholic acid, CA), and secondary BAs (deoxycholic acid, DCA) were generated for each sample, and each point represents a single sample. Box plots depict median and IQR, and statistics were performed using a Wilcoxon rank-sum, two-tailed, adjusted p-value.

**Figure S10:**
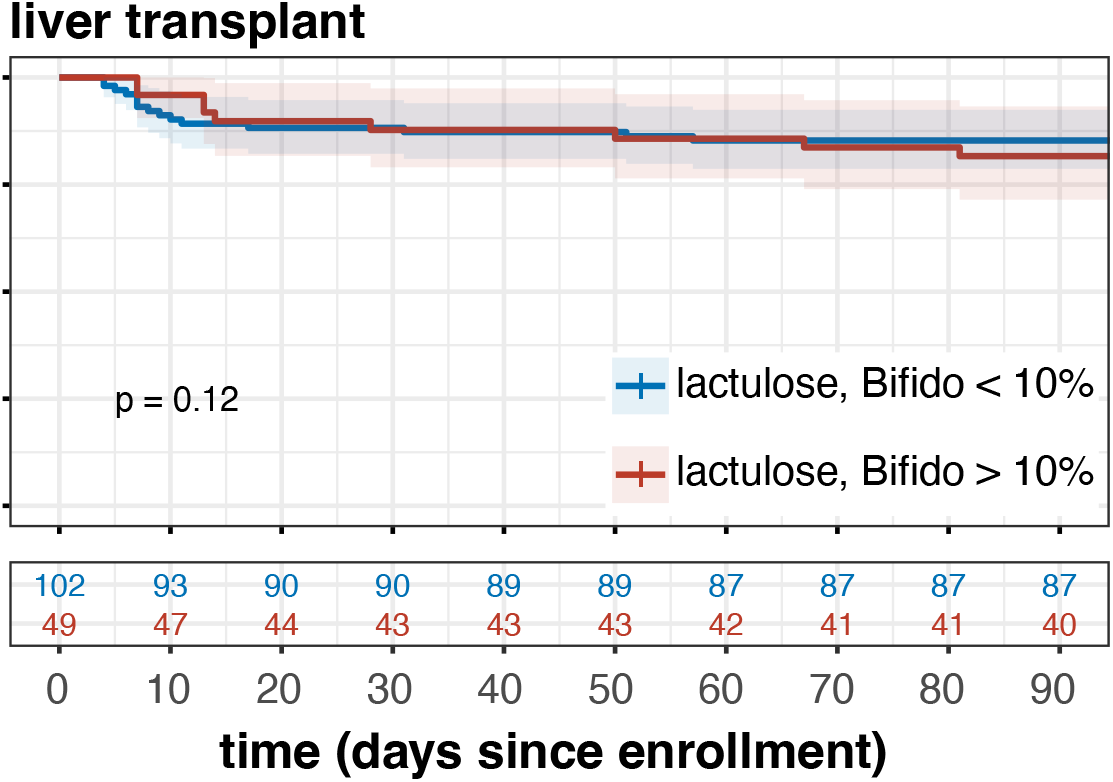
Initial sample characteristics relationship to liver transplant status. Kaplan-Meier curves were generated for liver transplant based on initial stool sample characteristics. Survival curves were stratified based on lactulose administration and *Bifidobacteria* expansion of the initial stool sample. The number at risk at each time-point is shown below.

**Table S1:** Ascites cell count and culture data are shown along with paired stool sample *Bifidobacterium, Enterococcus, Proteobacteria* (including *E. coli* and *Klebsiella*) abundance and collection time relative to ascites (negative indicating stool prior to ascites).

Taxon Relative Abundance (%)
| PMN | Ascites Culture | Stool-to-Ascites Time (days) | Bifidobacterium | Enterococcus | Proteobacteria | Escherichia coli | Klebsiella |
| --- | --- | --- | --- | --- | --- | --- | --- |
| 0 | GPC | 1 | 0.046 | 37.159 | 18.269 | 17.986 | 0.283 |
| 1139 | No growth | -11 | 0.000 | 95.524 | 0.000 | 0.000 | 0.000 |
| 7165.08 | No growth | 1 | 0.000 | 0.000 | 1.075 | 0.000 | 0.204 |
| 661.96 | No growth | 2 | 0.118 | 0.000 | 1.992 | 0.000 | 0.001 |
| 24050 | C. freundii, E. faecium | 1 | 0.600 | 5.677 | 6.383 | 0.000 | 1.536 |
| 4.895 | gram positive bacilli, C. jeikeium | 0 | 23.738 | 2.712 | 0.947 | 0.000 | 0.022 |
| 2105.1 | No growth | 0 | 0.130 | 86.176 | 1.709 | 1.696 | 0.000 |
| 1004.08 | No growth | -1 | 5.692 | 0.912 | 0.479 | 0.240 | 0.000 |
| 270 | No growth | -2 | 0.000 | 0.533 | 1.964 | 0.584 | 0.148 |
| 22847.5 | No growth | 0 | 0.000 | 0.000 | 0.000 | 0.000 | 0.000 |
| 1015.535 | No growth | 2 | 0.000 | 0.000 | 0.000 | 0.000 | 0.000 |
| 1585.08 | E. coli (ESBL) | 0 | 0.000 | 1.103 | 33.583 | 27.362 | 0.000 |
| 1085.4 | No growth | 2 | 9.024 | 2.340 | 15.366 | 15.312 | 0.000 |
| 7147.56 | P. multocida | 3 | 0.000 | 0.742 | 0.968 | 0.000 | 0.005 |
| 1730.52 | No growth | -3 | 0.000 | 89.197 | 0.003 | 0.000 | 0.000 |
| 1700.16 | C. albicans | 2 | 0.000 | 0.013 | 0.820 | 0.820 | 0.000 |
| 1549.24 | No growth | 0 | 0.000 | 0.016 | 8.967 | 8.750 | 0.000 |
| 77.665 | C. striatum | 2 | 0.000 | 100.000 | 0.000 | 0.000 | 0.000 |
| 790.72 | E. faecium | 1 | 0.776 | 13.923 | 69.340 | 68.771 | 0.275 |

**Table S2:** Blood culture data are shown along with paired stool sample *Bifidobacterium, Enterococcus, Proteobacteria* (including *E. coli* and *Klebsiella*) abundance and collection time relative to blood culture (negative indicating stool prior to culture).

Taxon Relative Abundance (%)
| Blood Culture | Stool-to-Blood Time (days) | Bifidobacterium | Enterococcus | Proteobacteria | Escherichia coli | Klebsiella |
| --- | --- | --- | --- | --- | --- | --- |
| E. faecium | 2 | 42.927 | 11.323 | 2.276 | 0.000 | 2.252 |
| S. marcescens | 4 | 8.824 | 82.094 | 0.280 | 0.000 | 0.000 |
| a-hemolytic streptococci | 0 | 0.000 | 0.000 | 1.075 | 0.000 | 0.204 |
| E. faecium | 4 | 0.000 | 95.907 | 0.000 | 0.000 | 0.000 |
| K. pneumoniae | 4 | 0.137 | 0.000 | 0.473 | 0.031 | 0.000 |
| P. aeruginosa | 0 | 0.000 | 99.091 | 0.134 | 0.000 | 0.000 |
| S. constellatus, Gemella species | 3 | 0.000 | 91.798 | 0.230 | 0.000 | 0.000 |
| K. pneumoniae (ESBL) | -3 | 0.000 | 73.261 | 7.628 | 0.000 | 7.628 |
| P. aeruginosa | 3 | 0.002 | 0.000 | 0.000 | 0.000 | 0.000 |
| K. pneumoniae (CRE) | 2 | 0.000 | 0.000 | 0.000 | 0.000 | 0.000 |
| E. coli (ESBL) | -3 | 0.000 | 0.000 | 40.989 | 27.224 | 0.000 |
| gram positive bacilli, M. abscessus | 2 | 0.000 | 10.450 | 51.974 | 45.901 | 0.191 |
| E. coli | 7 | 11.030 | 0.089 | 0.113 | 0.113 | 0.000 |
| S. aureus (MRSA) | 0 | 0.361 | 5.241 | 0.957 | 0.868 | 0.000 |
| K. pneumoniae | 2 | 0.945 | 0.013 | 1.000 | 0.871 | 0.108 |
| Bacillus species, not anthracis | 0 | 0.063 | 0.432 | 0.000 | 0.000 | 0.000 |
| E. coli | 3 | 0.000 | 8.036 | 9.342 | 9.342 | 0.000 |
| E. cloacae complex | 3 | 0.134 | 0.000 | 1.625 | 1.016 | 0.017 |
| coagulase negative Staphylococcus species, P. multocida | 3 | 0.000 | 0.742 | 0.968 | 0.000 | 0.005 |
| anaerobic gram positive bacilli | 5 | 2.840 | 1.701 | 3.106 | 0.006 | 0.000 |
| gram positive bacilli | 0 | 0.967 | 0.000 | 7.556 | 7.523 | 0.000 |
| S. hominis | 0 | 3.757 | 0.000 | 2.890 | 0.213 | 0.053 |
| C. kefyr | 1 | 0.000 | 31.960 | 24.890 | 24.554 | 0.015 |

## Notes

### Competing Interest Statement

The authors have declared no competing interest.

### Funding Statement

This work was funded privately by the Duchossois Family Institute at the University of Chicago
Authors were funded by the National Institutes of Health as follows: 
T32DK007074 (M.A.O.)
U01AA026975 (T.G.C.)

### Author Declarations

The Institutional Review Board of the University of Chicago gave ethical approval for this work.

